# Non-linear age dynamics of malaria infection and fine-scale environmental exposure in rural Uganda

**DOI:** 10.1101/2025.10.10.25337712

**Authors:** Max M. Lang, Violet Tuhaise, Phionah Kafuko, Aisha Nakato, Asmin Mohamed, Betty Nabatte, Narcis B. Kabatereine, Christl A. Donnelly, Goylette F. Chami

## Abstract

**Background:** Age-specific patterns of malaria are well-established for children aged ***<*** 5 years. Less understood is the epidemiology of malaria in older children and adults, and the influence of granular environmental risk.

**Methods:** We analyzed data from SchistoTrack, a community-based cohort in rural Uganda. We studied 4308 participants aged 5 to 90 years from 52 villages across three lakeside districts of Mayuge, Buliisa, and Pakwach, with enrollment between January 2022 to February 2024. The primary outcome was malaria infection status by rapid diagnostic test (RDT). Secondary outcomes included microscopy-confirmed infection with parasite density quantification and self-reported fever within the past month. We fitted a generalized additive mixed model (GAMM) with adaptive age smoothing, adjusting for sociodemographic factors, household characteristics, healthcare access, and environmental exposures. Environmental exposure was quantified using the Normalized Difference Vegetation Index (NDVI) derived from Sentinel-2 satellite imagery (10 m resolution), processed through hexagonal aggregation with Gaussian neighborhood smoothing and validated against field malacology surveys and participatory community mapping.

**Results:** Overall RDT prevalence was 41.2% (1776/4308), with microscopy prevalence at 32.3% (1363/4219), which was predominantly *Plasmodium falciparum* (83.1%; 1133/1363). Most infections were low-density (***<*** 999 parasites/µL; 71.6%; 976/1363). Malaria prevalence showed non-linear age patterns, peaking at 10 to 11 years then declining through adolescence before stabilizing in adulthood. Among RDT-positive individuals, fever prevalence decreased with age from 30.8% in children (aged 5 to 10 years) to 11.2% in adults (aged ***≥***20 years). Dense vegetation (per unit NDVI increase: Odds Ratio (OR) 3.25, 95% Confidence Interval (CI) 1.33–7.96) and greater distance from government health centers (per log-km: OR 1.87, 95% CI 1.34–2.59) increased the odds of infection. Proximity to vegetated water bodies increased the odds of infection compared to beaches: ponds/swamps (OR 1.65, 95% CI 1.19–2.28), river/river marsh (OR 1.63, 95% CI 1.16–2.31), lake marsh (OR 1.40, 95% CI 1.07–1.83).

**Conclusion:** Malaria prevalence remains high in older children and adults, though with fewer febrile cases, and is influenced by the local environment. Our findings support age-specific interventions targeting school-aged children while maintaining adult surveillance, and using validated environmental indices to guide sub-district resource allocation in high-risk areas.

## Introduction

Malaria remains a major cause of morbidity and mortality in sub-Saharan Africa (SSA) [1]. In 2022 alone, an estimated 249 million malaria cases occurred globally, with Uganda among the highest prevalence countries [1, 2]. Following successful control interventions, the burden has shifted from younger to older age groups, with school-aged children now showing the highest infection prevalence despite predominantly asymptomatic infections [3, 4]. However, how malaria risk changes across the age spectrum into adulthood remains underexplored, limiting evidence of age-infection relationships after accounting for environmental and sociodemographic determinants, which could potentially inform national programs.

Malaria surveillance and control programs in SSA have traditionally focused on children aged *<* 5 years, who account for most reported cases and severe outcomes [1, 2] with limited understanding of the epidemiology of older age groups. In the *<* 5 age group, infection prevalence typically increases from birth and peaks around 2 to 3 years before declining towards the age of 5 [1, 5]. School-aged children and adults have received less attention. However, recent studies indicate a shifting distribution of infections due to the widespread availability of interventions such as antimalarials and bed nets, showing high prevalence in older children and ongoing asymptomatic carriage in adults, both of which contribute to the reservoir for transmission [3, 6–9].

Local environmental conditions influence malaria epidemiology [10–12] and vector breeding is shaped by ecological features such as water type, vegetation density, and proximity to human dwellings [12–15]. Satellite-derived environmental indices, typically generated from reflectance data, have been widely incorporated into malaria models to map risk at different spatial scales, however primarily focusing on district or national analyses using 1 km *×* 1 km to 5 km *×* 5 km grids [16–19]. The increasing availability of freely accessible, high-resolution imagery, down to 10 m *×* 10 m allows investigation of environmental drivers at much finer scales [17, 20–22]. Yet, when used at this spatial resolution, such data present additional unresolved challenges, including pixel-level artifacts, grid-pattern distortions, and uncertainties about their correspondence with ground-truth measurements [23, 24]. Comparisons with field-based observations or qualitative approaches such as community mapping are often lacking, raising questions about the accuracy of such estimates despite these approaches being used in different contexts such as designing responses to environmental disasters [25–27]. While high-resolution satellite imagery is increasingly applied to detect and classify potential mosquito habitats and, more recently, incorporated into local malaria early warning systems [21, 28], its potential for mapping individual- and household-level risk remains underexplored [17]. Despite extensive control measures, including mass distribution of long-lasting insecticidal nets (LLINs) and periodic indoor residual spraying (IRS), many rural SSA communities continue to experience intense year-round transmission [29–32].

We conducted a large-scale study of 4308 individuals aged 5 to 90 years in rural Uganda. We assessed how malaria infection varies across age and identified relevant scales for environmental exposures derived from satellite data. Our aim was to investigate the non-linear age dynamics of malaria infection in older children and adults and to identify its determinants by investigating a wide range of sociodemographic, health access, and environmental risk factors.

## Methods

### Study design and participants

This study was conducted within SchistoTrack, a community-based prospective cohort in Uganda established in 2022 [33, 34]. The cohort is in three districts in Uganda: Pakwach along the River Nile, Buliisa along Lake Albert, and Mayuge along Lake Victoria. Households were randomly sampled using local village registers from the 52 total villages, beginning with 38 villages in 2022, the addition of 14 villages in 2023, and the inclusion of more households from existing villages in 2024. Households were eligible if they included at least one child and one adult who were residing in the community at least six months out of the year. At the end of the interview, the head of household designated one adult (aged *≥* 18 years) and one child (aged 5 to *<* 18 years) to undergo clinical evaluations using a paired sampling approach [34]. A total of 4321 individuals were clinically assessed at the time of recruitment between 2022 and 2024 [34]. Of these, 4308 participants from 2164 households obtained valid malaria test results (2022: 2877; 2023: 965; 2024: 466). The 13 individuals without malaria tests included those who attended clinical stations but did not complete the malaria testing station despite attending other clinical assessments. Of the 4308 participants who obtained a malaria test, 4219 (97.9%) had both RDT and microscopy results available for analysis.

### Malaria outcomes

The primary malaria outcomes were infection status and parasitemia intensity. Malaria infection status was determined using a RDT (Pan Ag Bioline Test) [35]. Infection status was coded as binary (positive/negative). Parasitemia intensity was assessed through thick and thin blood film slides following World Health Organization (WHO) guidelines for malaria microscopy [2, 36]. Parasitemia levels were classified as none, low (1–999 parasites/µL), moderate (1000–4999 parasites/µL), high (5000–99999 parasites/µL), and hyperparasitemia (*≥* 100000 parasites/µL). We collected data on fevers and malaria treatment through household questionnaires and clinical assessments. Fever was defined as self-reported fever within the past month. We also recorded self-reported use of antimalarial medication within the past month. Due to under-reporting during COVID-19, we restricted fever and treatment analyses to 2023–2024 data (*n* = 1431). There were only two fever reports, both RDT-negative, among 2877 participants enrolled in 2022. Village-level bed net coverage was calculated from self-reported use, but only available from 2023 onward. Additional details are provided in the supplement (see S1 Text).

### Environmental exposure

We analyzed Sentinel-2 Level-2A surface reflectance imagery (10 m resolution) from December-January periods (2021–2024) accessed through Google Earth Engine, filtered for *<*20% cloud cover. We calculated Normalized Difference Vegetation Index (NDVI), Normalized Difference Water Index (NDWI), Modified Normalized Difference Water Index (MNDWI), and Enhanced Vegetation Index (EVI) following standard formulations [37]. To reduce noise while preserving spatial detail, we applied hexagonal grid aggregation (*≈*900 m^2^ cells) with Gaussian-weighted neighborhood smoothing. Environmental indices were validated against 437 malacology surveys documenting water body characteristics and participatory community mapping exercises. Community mapping involved participatory exercises during fieldwork in 2025, where village residents created hand-drawn maps identifying key environmental features, water access points, and areas of perceived malaria risk. For validation, we compared satellite-derived indices against 437 ground-truth malacology surveys and these community maps. We assessed the ability of each index to discriminate between ecological scenarios (e.g., beach vs. non-beach sites) and examined correlations with field-recorded vegetation counts. Based on validation analyses, we selected smoothed NDVI as the primary environmental predictor for modeling. The full approach is described in the supplement (see S3 Text: Environmental Data Processing). Water body type of the nearest water site (categorical; beach, marsh, river, pond, or rice paddy, see Iacovidou et al. [38]) was included to represent broader ecological features beyond the immediate household environment that influence mosquito breeding suitability.

### Sociodemographic and health access covariates

Household interviews included individual-, household-, and village-level information [34, 39]. Individual characteristics of age (years), gender (female as reference), years of formal education (0–14), and occupation (categorical; none/other as reference, including students, unemployed, and farmer, fisherman, or fishmonger based on main income-earning activity). For each individual, we determined whether they belonged to the majority tribe or majority religion within their village (using a simple majority and coded as binary) [34, 39]. Household socioeconomic indicators included a composite home-quality score (3–12 scale) based on wall material, roof type, and floor quality, with higher scores indicating better construction [39]. We recorded electricity access (grid or solar), home ownership, number of rooms, and household size. Household social status was coded as positive if any member held a recognized local leadership position. Water, sanitation, and hygiene (WASH) indicators were recorded at the household level and included hand-washing facilities with water and soap availability, sanitation facility type (improved versus unimproved per WHO / United Nations International Children’s Emergency Fund (UNICEF) criteria), any water treatment practices, and access to improved water sources [40]. Health facility locations were obtained through field surveys (2022–2023) and OpenStreetMap data. We computed Euclidean distances from each household to the nearest government health center and drug shop, with distances log-transformed as *log*_10_(1 +distance in kilometers) [34]. District (Mayuge, Buliisa, Pakwach) and recruitment year (2022–2024) were included to account for the study design of the SchistoTrack cohort.

### Statistical Analysis

Data analysis was completed in R version 4.2.1. We excluded individuals with invalid or missing RDT results from all analyses. For comparisons between groups, we used Pearson’s chi-squared tests for categorical variables and Wilcoxon rank-sum tests for continuous variables. Spearman’s rank correlation also was used for all correlation analyses. All statistical tests were two-sided with significance set at *α* = 0.05.

To assess spatial clustering of malaria cases, we aggregated data to the household level, classifying households as positive if either sampled individual tested positive. We calculated Moran’s *I* statistic across a range of *k*-nearest neighbor specifications (*k* = 2 to 50) with row-standardized spatial weights, implemented using the spdep package.

Prior to modeling, we examined pairwise Spearman’s correlations among all candidate predictors, excluding variables with *|r| >* 0.6 to avoid multicollinearity. We fitted a logistic Generalized Additive Mixed Model (GAMM) using the mgcv package to capture non-linear relationships between age and malaria risk while accounting for village-level clustering:

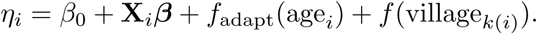

The age effect was modeled using an adaptive smoothing spline with *k* = 20 basis functions and an order-3 penalty basis (*m* = 3). Village-specific random effects were included. Parameters were estimated via penalized Restricted Maximum Likelihood (REML). Model performance was evaluated through 10-fold cross-validation with Receiver operating characteristic (ROC) curve analysis. We conducted sensitivity analyses including restricting to *P. falciparum* monoinfections and comparing smoothed versus unsmoothed NDVI values. Complete model specifications and mathematical details are provided in the supplement (see S4 Text).

## Results

### Malaria status and parasitemia

Overall malaria prevalence diagnosed via RDT was 41.2% (1776/4308). The malaria prevalence diagnosed via microscopy was 32.3% (1363/4219) of which 83.1% (1133/1363) were *P. falciparum* monoinfections, 1.1% (15/1363) were *P. malariae* monoinfections, and 15.8% (215/1363) were mixed infections where nearly all mixed infections included *P. malariae* and *P. falciparum* (214/215). There were no monoinfections of *P. vivax* or *P. ovale*, although these species were detected in one mixed infection. Among the 1363 microscopy-positive cases with confirmed parasites, parasitemia was predominantly low. The geometric mean across all positives was low at 454 parasites/µL (95% CI 416–497). The vast majority were low-density infections (1–999 parasites/µL; 976/1363, 71.6%), about one-fifth were moderate (1000–4999 parasites/µL; 257/1363, 18.9%). High parasitemia (5000–99999 parasites/µL) was observed in 128/1363 (9.4%), while hyperparasitemia (*≥* 100000 parasites/µL) was rare (2/1363, 0.1%).

Among the subset of participants enrolled in 2022 and followed through 2024 (*n* = 2887), transitions between parasitemia categories were mostly movement from low-density infection to no parasites (see Figure S14). Village-level bed net coverage improved between 2023 and 2024. In Pakwach, coverage increased significantly from 84.6% to 96.0% (*χ*^2^ = 15.645, *p <* 0.001), while coverage remained consistently high (*>*93%) in Buliisa and Mayuge throughout the study period.

The relationship between malaria infection and clinical symptoms varied by age group, based on data from 2023–2024 (Table 1). The age of participants ranged from 5 to 90 years. Median age was 18 years (Interquartile range (IQR) 9–38), with 54.5% (2346/4308) female participants. Adults aged *≥* 60 years accounted for 5.6% of the sample (240/4308). Among children aged 5 to 10 years, 30.8% (81/263) of RDT-positive cases reported fever symptoms compared to 16.3% (27/166) of RDT-negative children. This proportion decreased with age. In adolescents aged 11 to 19 years, 21.9% (42/192) of RDT-positive individuals reported fever versus 15.8% (16/101) of RDT-negative individuals. There were fewer symptomatic infections in adults than in children. Adults aged *≥* 20 years showed the lowest proportion of febrile infections, with 11.2% (20/179) of RDT-positive adults reporting fever compared to 5.8% (31/530) of RDT-negative adults. Across age groups for 2023–2024, 22.6% (143/634) of RDT-positive individuals reported fever symptoms while 77.4% (491/634) did not (Table 3). There was a significant association between self-reported fever in the past month and RDT positivity (*χ*^2^ = 48.34, *p <* 0.001). Antimalarial use within the past one month was common among participants from 2023–2024 and showed a significant age-dependent trend, with the highest treatment rates in children aged 5 to 10 years (54.8%), followed by adolescents (49.5%), and adults (38.4%; *χ*^2^ = 50.04, *p <* 0.001). For RDT-positive individuals only, we found no statistically significant difference in fever prevalence between those who reported recent antimalarial use and those who did not for any age group (Table S4).

**Table 1:** Self-reported fever prevalence by malaria RDT status across three age groups (5 to *<* 11, 11 to *<* 20, and ≥ 20 years) among participants recruited in 2023–2024, showing age-related decline in symptomatic infections.

| Fever Status | Age Group | RDT Positive |  | RDT Negative |  |
| --- | --- | --- | --- | --- | --- |
|  |  | n | (%) | n | (%) |
| Fever reported | 5 to $< 11$ years | 81 | 30.8 | 27 | 16.3 |
| No report | 5 to $< 11$ years | 182 | 69.2 | 139 | 83.7 |
| **Total** | 5 to $< 11$ years | 263 | 100.0 | 166 | 100.0 |
| Fever reported | 11 to $< 20$ years | 42 | 21.9 | 16 | 15.8 |
| No report | 11 to $< 20$ years | 150 | 78.1 | 85 | 84.2 |
| **Total** | 11 to $< 20$ years | 192 | 100.0 | 101 | 100.0 |
| Fever reported | $\geq 20$ years | 20 | 11.2 | 31 | 5.8 |
| No report | $\geq 20$ years | 159 | 88.8 | 499 | 94.2 |
| **Total** | $\geq 20$ years | 179 | 100.0 | 530 | 100.0 |

**Table 2:**
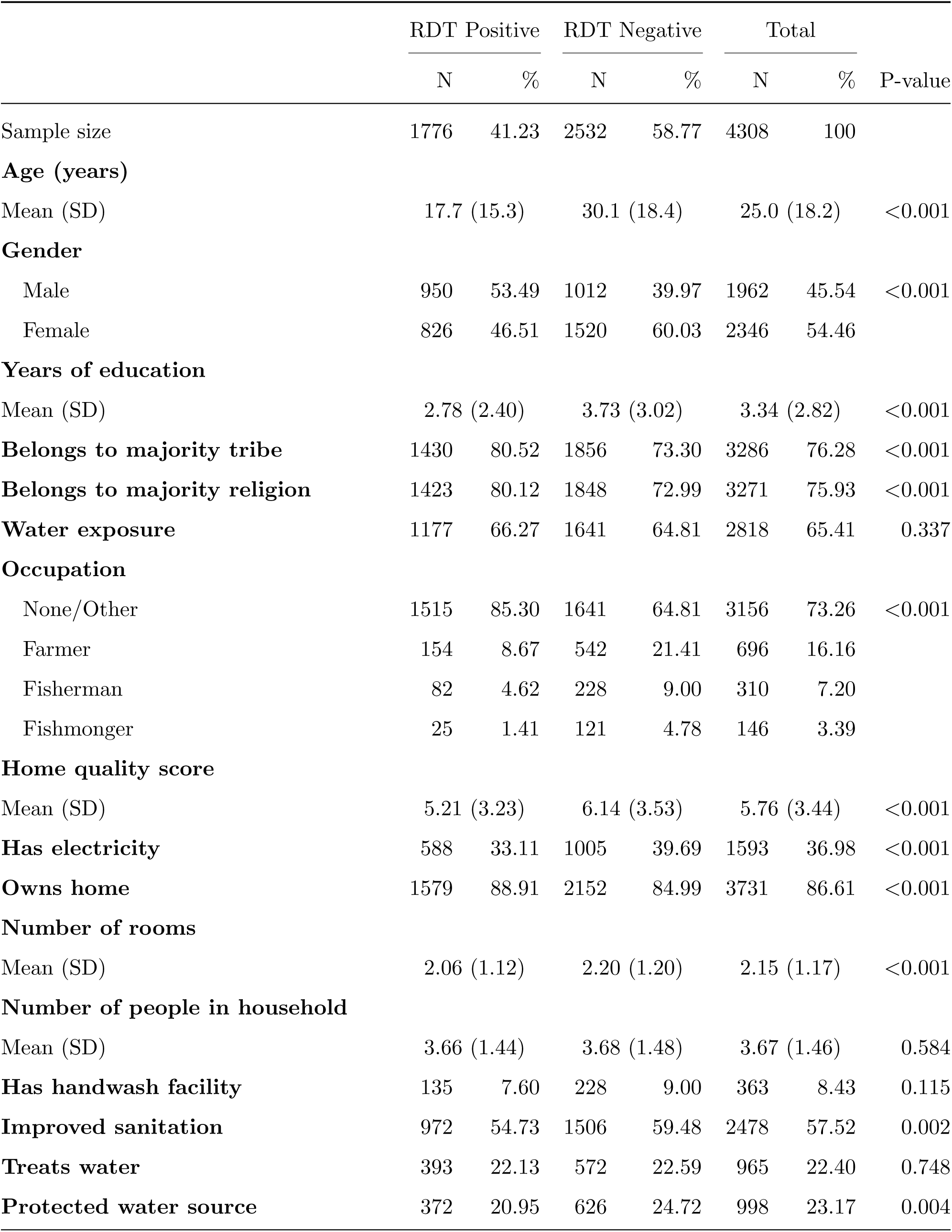
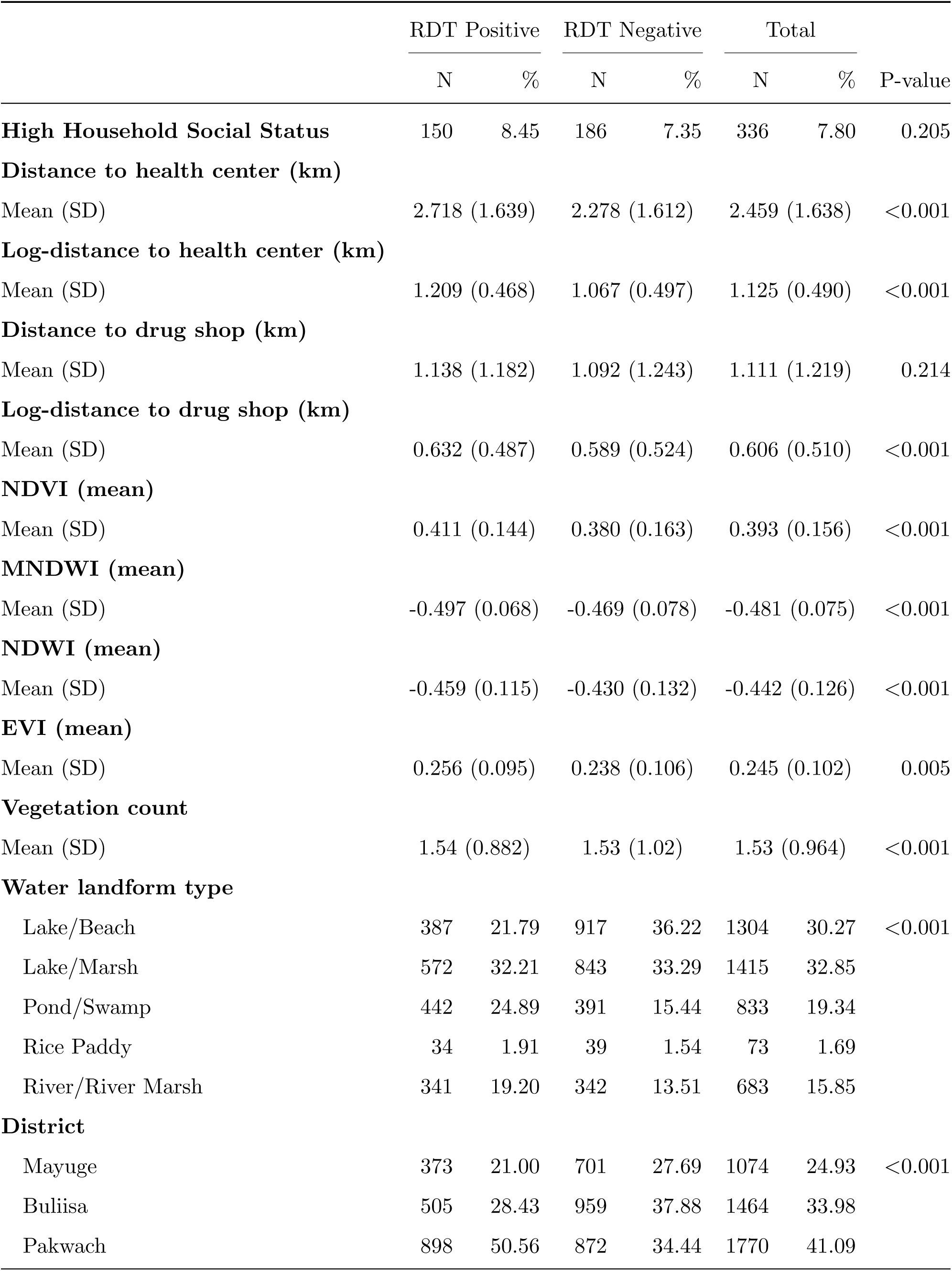
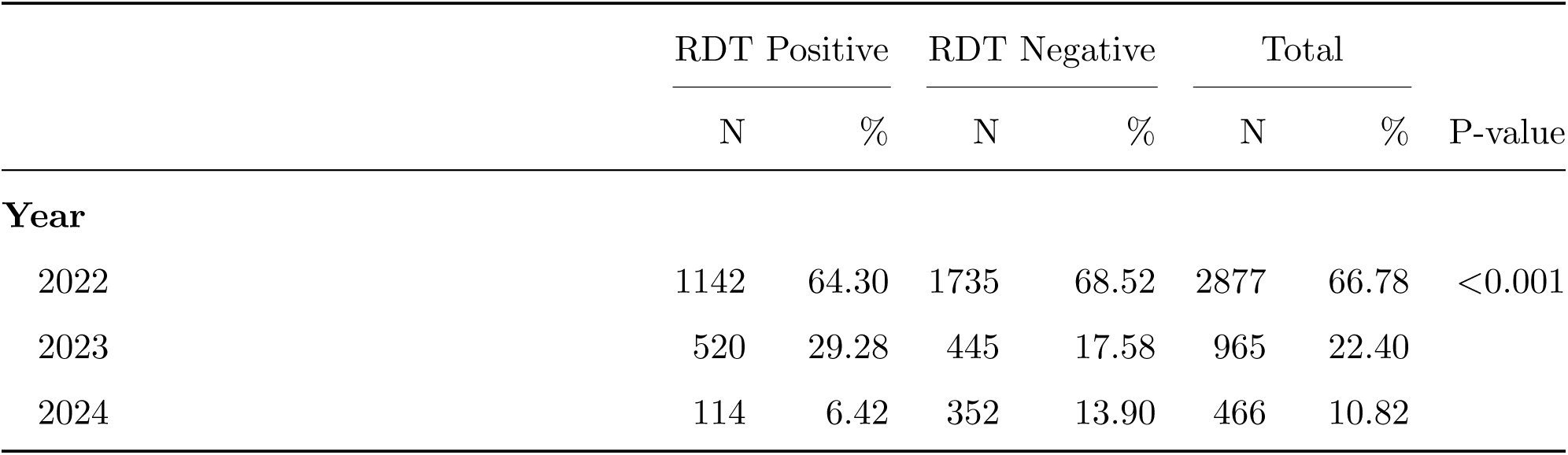
Comparison of sociodemographic, environmental, and clinical characteristics of study participants, stratified by Rapid Diagnostic Test (RDT) results (N=4308).

**Table 3:** Proportion of participants reporting fever within the past month by malaria RDT status among 1431 individuals from 2023–2024, 2022 data excluded due to under-reporting of COVID-19-related symptoms.

| Fever Status | RDT Positive |  | RDT Negative |  |
| --- | --- | --- | --- | --- |
|  | n | (%) | n | (%) |
| Fever reported | 143 | 22.6 | 74 | 9.3 |
| No report | 491 | 77.4 | 723 | 90.7 |
| **Total** | 634 | 100.0 | 797 | 100.0 |

A summary of participant sociodemographic, environmental, and clinical characteristics stratified by RDT status is provided in Table 2. 73.3% (3156/4308) participants had no formal occupation or other activities (including students and unemployed), 16.2% (696/4308) were farmers, 7.2% (310/4308) were fishermen, and 3.4% (146/4308) were fishmongers. The most common tribes were Alur (61.1%, 2632/4308), Musoga (12.2%, 527/4308), and Bagungu (9.4%, 407/4308). Religious affiliations included Christians (73.9%, 3184/4308), Islam (12.8%, 552/4308), and Born-Again Christians (12.0%, 519/4308). Almost 30% of participants lived in single-room households with more than 3 people (see Table S3).

### Evaluation of environmental exposure indices

The processed Sentinel-2 imagery yielded two vegetation and two water indices aggregated to hexagonal grids (*≈*900 m^2^ cells) using Gaussian-weighted smoothing with first- and second-order neighbors. The spatial smoothing process improved data quality (Figure 1c,d). Raw NDVI values exhibited high variability and discontinuous patches, especially along and within water-land interfaces. The Gaussian-weighted smoothing reduced these spatial artifacts while preserving ecological features, such as shorelines as well as areas with low vegetation. Environmental indices were strongly correlated (NDVI–EVI: *ρ* = 0.986, *p <* 0.001; NDVI–NDWI: *ρ* = *−*0.983, *p <* 0.001). For household locations (*n* = 2164), NDVI values differed across districts (Table 4), ranging from a median of 0.291 in Buliisa to 0.520 in Mayuge, and the IQRs demonstrated variation within each district (Buliisa: 0.167, Pakwach: 0.168, and Mayuge: 0.182) to capture local variability in environmental features.

**Fig. 1:**
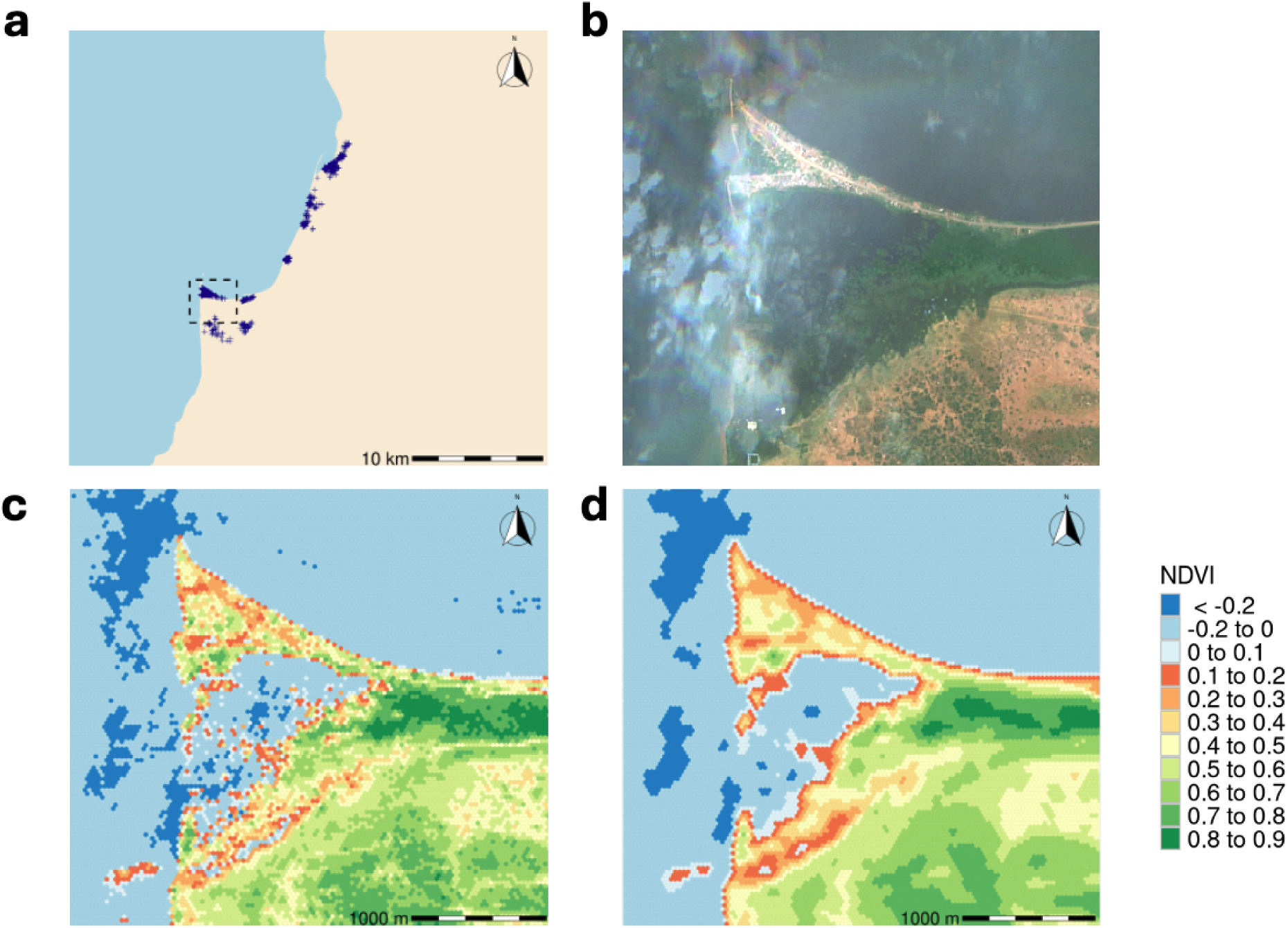
Spatial distribution of study sites and NDVI analysis. **(a)** Regional map showing household locations (blue markers). **(b)** Sentinel-2 satellite imagery of the area outlined in (a). **(c)** Unsmoothed NDVI values. **(d)** Spatially smoothed NDVI used for statistical analysis. The color gradient in (c) and (d) indicates vegetation density, from low (red) to high (green).

**Fig. 2:**
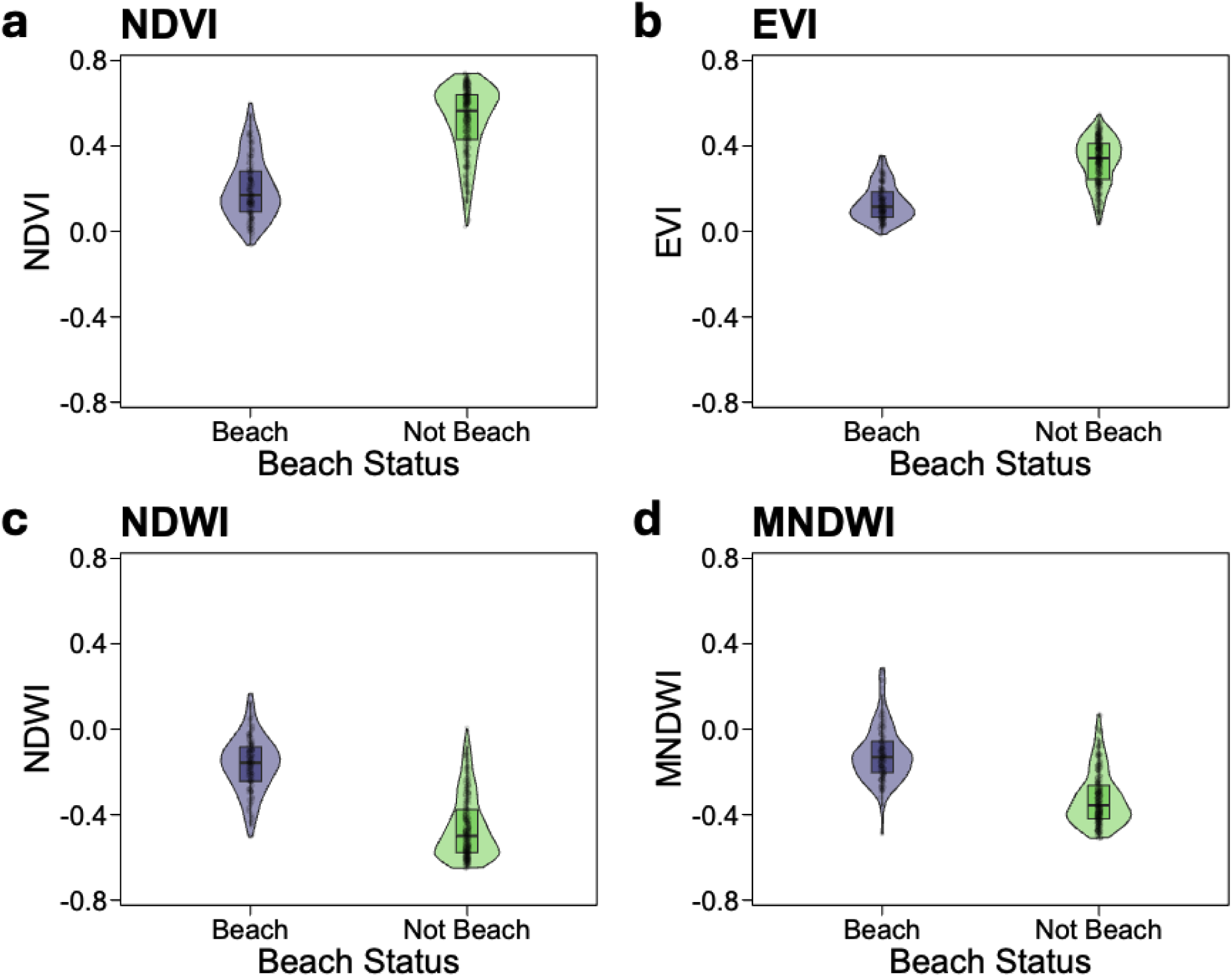
Validation of four satellite-derived environmental indices using ground truth data from a malacology survey. The boxplots compare the distribution of values for **(a)** NDVI, **(b)** EVI, **(c)** NDWI, and **(d)** MNDWI between survey sites identified on the ground as either ‘beach’ or ‘non-beach’.

**Table 4:**
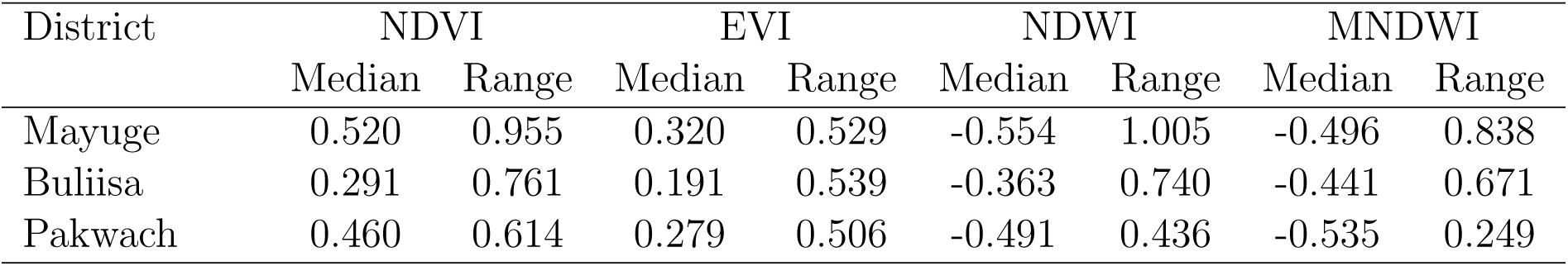
Environmental indices for household locations by district.

| District | NDVI |  | EVI |  | NDWI |  | MNDWI |  |
| --- | --- | --- | --- | --- | --- | --- | --- | --- |
|  | Median | Range | Median | Range | Median | Range | Median | Range |
| Mayuge | 0.520 | 0.955 | 0.320 | 0.529 | -0.554 | 1.005 | -0.496 | 0.838 |
| Buliisa | 0.291 | 0.761 | 0.191 | 0.539 | -0.363 | 0.740 | -0.441 | 0.671 |
| Pakwach | 0.460 | 0.614 | 0.279 | 0.506 | -0.491 | 0.436 | -0.535 | 0.249 |

NDVI showed the best discrimination between scenarios, particularly for beach habitats where it achieved a rank-biserial correlation of *ρ* = *−*0.833 (*p <* 0.001). Beach locations showed lower vegetation coverage (median NDVI: 0.17, IQR: 0.09–0.28) compared to non-beach sites (median: 0.51, IQR: 0.40–0.63). While water indices (NDWI, MNDWI) also discriminated beach habitats effectively (*ρ* = 0.835 and 0.761 respectively), NDVI consistently provided strong differentiation across all ecological scenarios including marsh habitats (*ρ* = 0.458, *p <* 0.001) and sites with mosquito-suitable floating vegetation (*ρ* = 0.432, *p <* 0.001; see Supplementary Figure S10, Figure S11, and Figure S16). NDVI also was robust when compared to community mapping. Figure 3 shows that areas marked as roads and village centers with trading centers on community maps corresponded to zones of lower NDVI values (shown in yellow/orange), reflecting reduced vegetation in these areas.

**Fig. 3:**
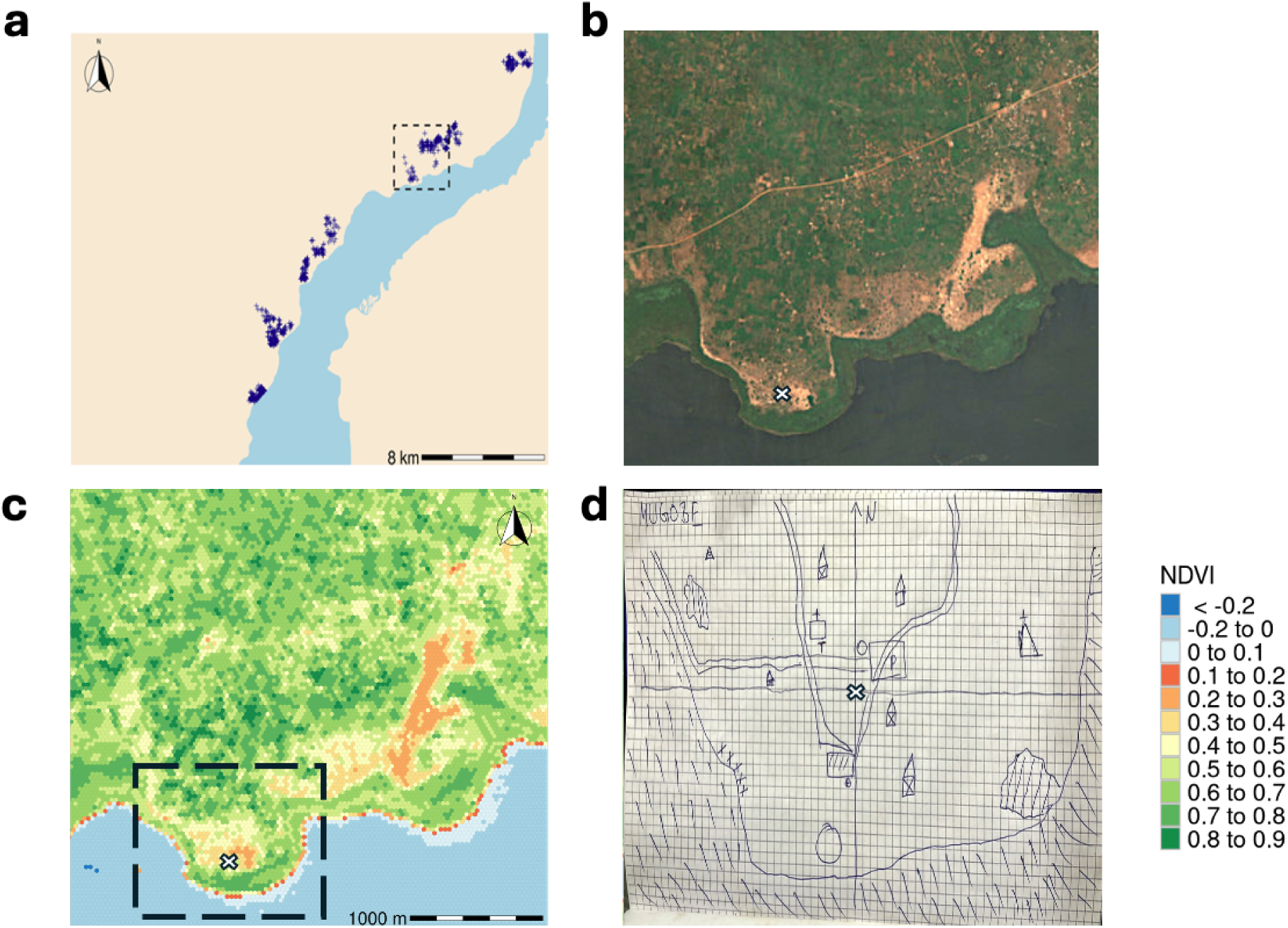
Comparison of satellite-derived NDVI with a community participatory map from 2025. **(a)** Regional overview. **(b)** Satellite imagery. **(c)** Spatially smoothed NDVI. **(d)** Community participatory map. The white cross marks the center and the square box approximates the area depicted in the community map **(d)**. The participatory map illustrates local perceptions of environmental features, water bodies, and malaria risk zones as identified by residents.

### Non-linear age dynamics of malaria infection

Generalized additive modeling confirmed an age-dependent pattern, with the smoothed prevalence curve reaching maximum values at 11 years (Figure 5b). The smooth function required 6.8 effective degrees of freedom, indicating substantial deviation from a linear relationship. After adjustment for covariates in the full model (Figure 4), peak prevalence occurred slightly earlier at 10.1 years (Figure 5c). The adaptive smoothing spline for age allowed a non-linear relationship with malaria risk (effective degrees of freedom = 6.8, *p <* 0.001). The instantaneous rate of change shows the age effects across the lifespan (Figure 5d) on the log-odds scale. For example, at age 12.7 years—identified as a point of locally minimal curvature through examination of the second derivative—the relationship between age and log-odds of infection was approximately linear. At this specific age, we interpreted the instantaneous rate of change as approximating a one-year increase. The odds ratio was 0.92 (95% CI 0.89–0.95), representing an 8% decrease in the odds of infection per increase in each year of age, however, this interpretation was only valid near this inflection point. At ages where the curve showed high curvature (such as ages 7–10 or 15–20), the effect of a one-year age increase differed from the instantaneous rate of change. This age-prevalence relationship contrasted with parasitemia density patterns, where geometric mean parasite loads peaked among individuals aged five years rather than pre-adolescents (Figure 6).

**Fig. 4:**
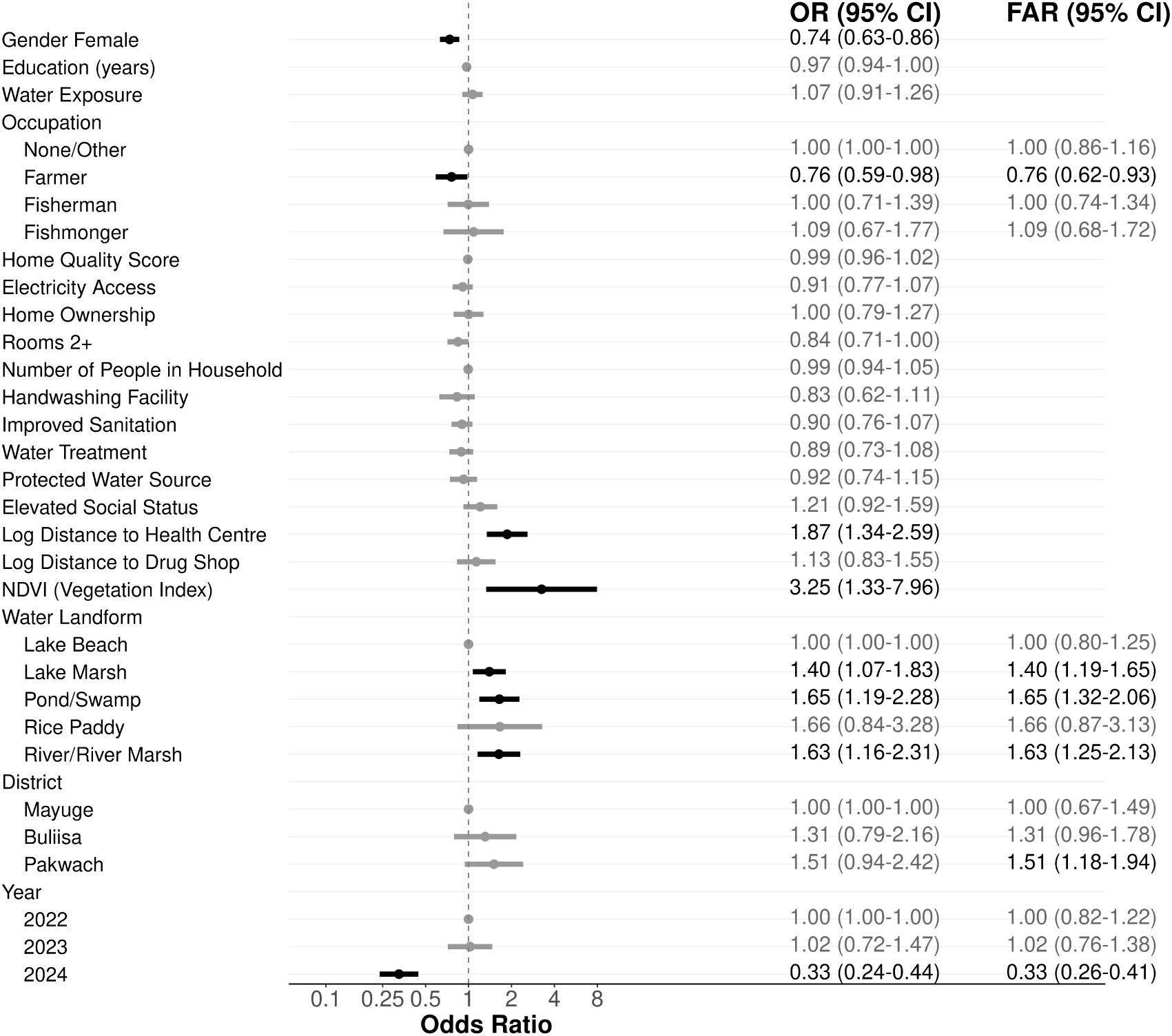
Determinants of malaria infection. The forest plot shows adjusted odds ratios and 95% confidence intervals for predictors of malaria infection status from the final GAMM.

**Fig. 5:**
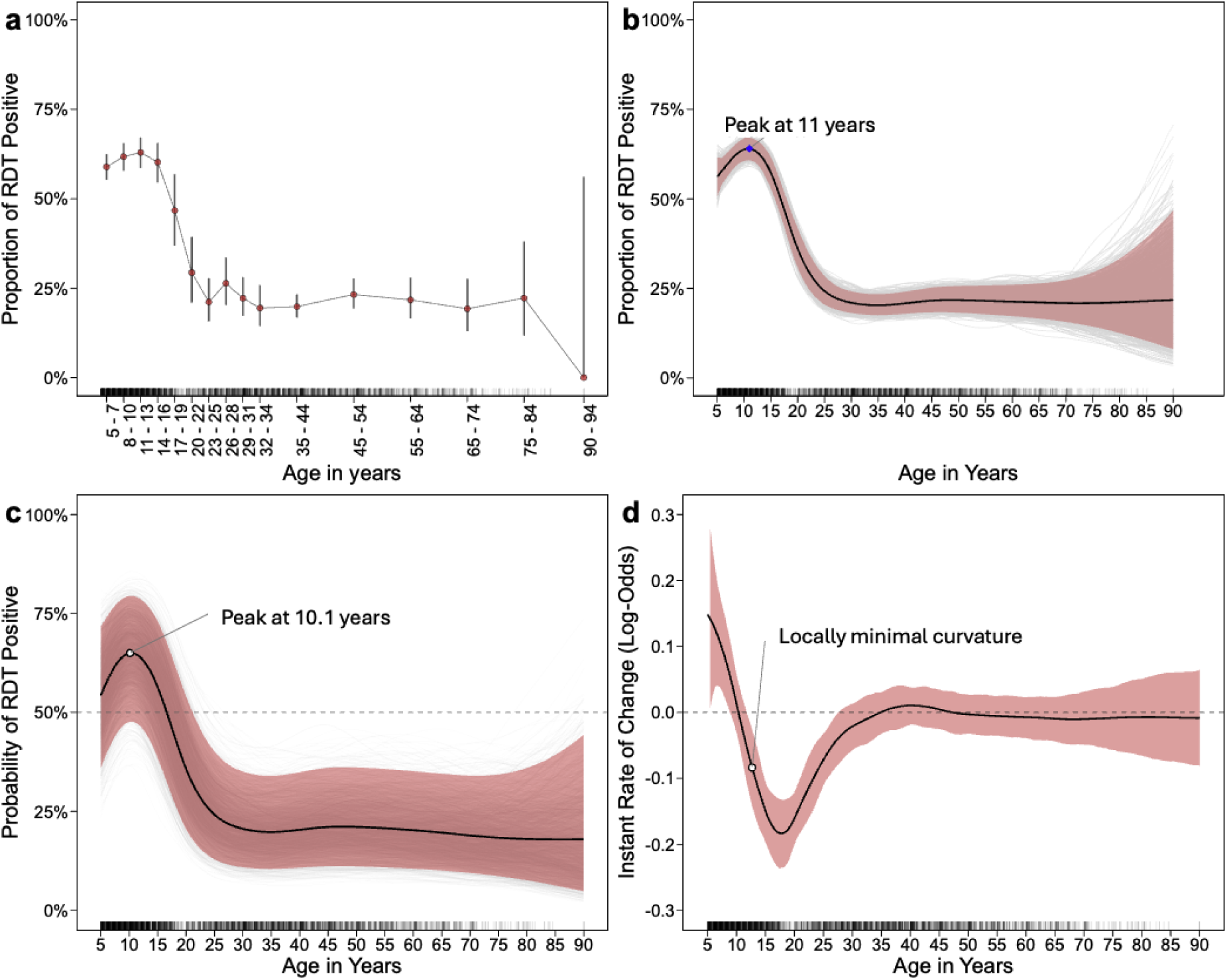
Age-specific patterns of malaria infection. **(a)** Proportion of individuals with malaria infection by age with 95% confidence intervals. **(b)** Smoothed age-infection relationship from the GAMM with 95% confidence bands. **(c)** Adjusted age-infection relationship from final GAMM with 95% confidence bands. **(d)** First derivative of age effect on log-odds Scale.

**Fig. 6:**
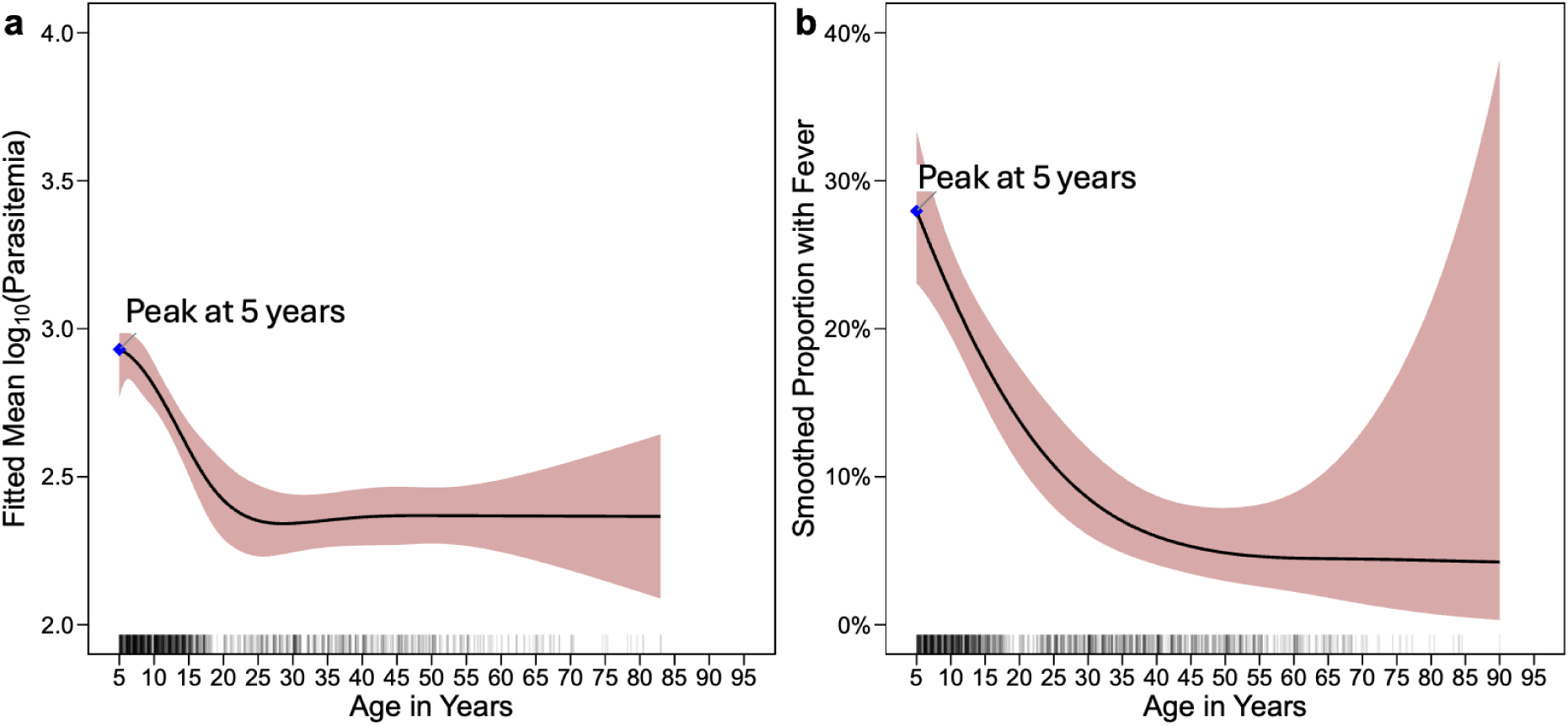
(a) Fitted mean log_10_ parasitemia among microscopy-positive infections by age; solid line from a GAMM with 95% confidence band. Parasitemia peaks at 5 years and declines through adolescence, remaining low in adults. **(b)** Smoothed proportion of RDT-positive participants reporting fever in the past month (2023–2024 subset of individuals) versus age, with 95% confidence band. The symptomatic fraction is highest around 5 years and falls sharply with age, reaching low levels in adulthood. Rug plots show the age distribution of observations.

### Sociodemographic, environmental, and health access determinants of malaria

Figure 4 shows the malaria infection model and results for covariates other than age. Gender was significantly associated with infection, with females having 0.26 lower odds than males (OR 0.74, 95% CI 0.63–0.86, *p <* 0.001). Farmers had similarly lower odds compared to individuals without an occupation (OR 0.76, 95% CI 0.59–0.98, *p* = 0.037), while fishermen showed no significant difference compared to individuals with no occupation (OR 1.00, 95% CI 0.71–1.39, *p* = 0.966). Environmental factors showed strong associations as each one-unit increase in NDVI corresponded to 3.25-fold increase in odds of infection (OR 3.25, 95% CI 1.33–7.96, *p* = 0.006). NDVI is normalized and bounded in [*−*1, 1], such that a one-unit increase is relatively extreme, thus, considering per 0.10 NDVI, we observed a 1.12-fold increase in odds. Further, based on the medians obtained through our ground-truth data (beach NDVI = 0.17 vs. non-beach = 0.51; see Figure 2), moving from a beach-like to a non-beach value (Δ = 0.34) corresponds to a 1.50-fold increase in odds (holding other covariates fixed). Proximity to specific water-body types with vegetation suitable for mosquitoes increased odds as compared to beaches. Ponds/swamps had the highest odds (OR 1.65, 95% CI 1.19–2.28, *p <* 0.001), followed by rice paddies (OR 1.66, 95% CI 0.84–3.28, *p* = 0.099), river/river marsh (OR 1.63, 95% CI 1.16–2.31, *p* = 0.004), and lake marshes (OR 1.40, 95% CI 1.07–1.83, *p* = 0.009). The nearest water-landform covariate contributes information at a spatial scale that was not redundant with the immediate (32–64 m) NDVI smoothing around households. 2.7% (117/4308) of participants lived within 32.2 m of their nearest water site, while 10.5% (452/4308) lived within 64.4 m.

Each one-unit increase in log-distance to the nearest health center was associated with a 1.87-fold increase in the odds of infection (OR 1.87, 95% CI 1.34–2.59, *p* = 0.003). For illustration, comparing households 5 km to 1 km from a health center under log_10_(1 +distance in kilometers) gives an 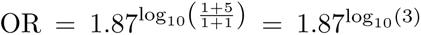 *≈* 1.35-fold increase. An individual recruited in 2024 had lower odds of having malaria than individuals recruited in 2022 (OR = 0.33, 95% CI 0.24–0.44, *p <* 0.001).

### Model performance and sensitivity analyses

For the malaria infection status diagnosed via RDT, the model explained 21.9% of the total deviance. The inclusion of a village-level random effect in our final GAMM accounted for spatial dependency observed in the data, as evidenced by the reduction in spatial autocorrelation from Moran’s *I* = 0.025 (*p <* 0.001) in standard GAMM residuals to Moran’s *I* = −0.003 (*p* = 0.574) in the final model. 10-fold cross-validation yielded an area under the ROC curve of 0.787 (Figure S17). Model performance was further evaluated using a confusion matrix with associated metrics including Cohen’s kappa coefficient for agreement beyond chance see Table S6). Using the optimal classification threshold of 0.399 determined by the Youden Index, the model achieved 72.92% sensitivity and 71.37% specificity (Positive Predictive Value (PPV) = 64.11%, Negative Predictive Value (NPV) = 78.98%). The model correctly classified 72.01% of observations overall (95% CI: 70.64%–73.34%). The complete cross-validated confusion matrix and associated performance metrics are provided in the supplement Table S6.

Our findings remained robust across multiple sensitivity analyses. Restricting the analysis to *P. falciparum* monoinfections (*n* = 1376) yielded similar associations for most predictors, though the NDVI effect with fewer observations in the model was attenuated and no longer significant (OR 1.82, 95% CI 0.74–4.50, *p* = 0.185). The expanded case definition incorporating antimalarial treatment recipients as a positive malaria case showed consistent patterns except for temporal trends, where participants enrolled in 2023 demonstrated higher odds than participants enrolled in 2022 (OR 3.24, 95% CI 2.34–4.49, *p <* 0.001). Alternative vegetation indices performed similarly to NDVI, with EVI showing comparable effect sizes (OR 6.37, 95% CI 1.97–20.62, *p* = 0.005), while water indices (NDWI, MNDWI) demonstrated expected inverse relationships with infection (NDWI: OR 0.14, 95% CI 0.04–0.51, *p* = 0.001; MNDWI: OR 0.09, 95% CI 0.01–0.46, *p* = 0.008). Spatial smoothing of NDVI values produced a stronger and significant association with malaria risk (OR 3.25, 95% CI 1.33–7.96, *p* = 0.006) when compared to the weaker, insignificant effect size of unsmoothed NDVI (OR 1.78, 95% CI 0.90–3.53, *p* = 0.1) (Figure S15).

## Discussion

A better understanding of determinants of malaria infection and the clinical relevance of current infection is essential for identifying high transmission areas and tailoring interventions in resource-limited settings. We investigated the relationship between malaria infection with a comprehensive set of sociodemographic, health access, and environmental variables in older children and adults within the SchistoTrack cohort, and developed a framework for validating high-resolution satellite-derived environmental indices to identify risk areas at both district and village scales. We showed a high malaria prevalence in Ugandan lakeside communities and identified a non-linear age-infection relationship characterized by peak prevalence at 10 to 11 years.

Malaria was age-dependent and non-linear for individuals aged 5 years and older. The nonlinearity of this relationship was most evident between ages 5 to 20 years, where prevalence increased steadily from age five to peak at 10 to 11 years, then declined sharply through adolescence before plateauing in adulthood. Our findings contribute to growing evidence that school-aged children both maintain high susceptibility to malaria and serve as a critical reservoir for community-wide transmission, with infection prevalence peaking at age 10 to 11 years and most infections remaining asymptomatic, yet perpetuating the transmission cycle across all age groups [3, 4, 8, 9, 30, 41, 42].

We observed a steep increase in risk from ages 5 to 10 years. This age group has historically received less attention in malaria control programs, with current WHO guidelines providing only a conditional recommendation for intermittent preventive treatment in school-aged children [2]. The steep decline after age 10 could potentially reflect the gradual acquisition of immunity through repeated exposure [43]. Longitudinal studies documented variation in how children develop immunity to malaria [43]. Some children stop experiencing clinical malaria episodes relatively early, yet continue to harbor asymptomatic infections with high parasite densities and maintain elevated antibody levels. This pattern suggests that clinical immunity can develop independently of parasitological immunity. However, early chemoprophylaxis may interfere with this natural immunity acquisition process [44]. There are also behavioral or cultural factors that may explain why children have increasing rates of malaria prevalence after five years of age. After five years of age, children experience increased outdoor activity and less parental supervision [45, 46], engaging in unsupervised outdoor activities during peak mosquito-biting hours [46, 47].

In addition to the non-linear malaria prevalence curves, we showed that children aged 5 to 9 years have high levels of parasitemia and commonly fever associated with malaria-positive RDTs. Despite pre-adolescents showing peak infection prevalence, febrile malaria cases were most common in individuals aged 5 years (30.8%) and declined with age, as only 11.2% of individuals aged *≥* 20 years had febrile malaria. Despite the prevalence being consistent in adults with no change from years 25 and older, the burden of malaria was still high; approximately one in five adults (25 to 90 years) tested RDT positive. Adults had predominantly asymptomatic low-density infections. School-aged children are becoming an increased focus of trials [4, 42, 48]. Given the burden in school-aged children and the generally high malaria prevalence in our study population, school-based intermittent preventive treatment may be appropriate [4, 8, 42], though implementation could potentially lead to further age distribution shifts as already observed in some settings [3] and might face challenges, such as poor school attendance and community trust issues [4]. Further, asymptomatic infections in adults are increasingly being recognized as important contributors to community transmission, as demonstrated in other endemic settings [3, 4, 8, 9, 41, 49, 50]. These findings suggest that effective malaria control requires age-stratified interventions focusing on school-based programs targeting the peak transmission age group alongside continued surveillance and treatment of the asymptomatic adult reservoir. The high proportion of individuals reporting recent antimalarials suggests that the overall malaria prevalence of 41.2% may be a conservative estimate of the true infection burden. Widespread treatment could clear infections before detection, potentially masking the full extent of transmission. However, because the highest rates of treatment were observed in younger age groups, the age-related trends we identified are unlikely to be altered. Furthermore, to ensure our findings on symptoms were robust, we compared fever prevalence among treated and untreated RDT-positive individuals. We found no significant difference in any age group, suggesting the high burden of asymptomatic infection is not an artifact of recent treatment.

Gender influenced the likelihood of infection. Males showed higher odds of infection compared to females with an effect that persisted even when restricting analysis to individuals aged *<* 20 years. Women often maintain closer proximity to households and may have different healthcare-seeking behaviors than men in rural SSA settings [51]. Evidence from Pullan et al. [52] suggests that adult males are least likely to use bed nets and only 27% of school-aged children sleep under a net, while usage was highest among children *<* 5 years old and women of reproductive age [52]. This difference in gender-specific prevalence (as well as age-specific prevalence) could potentially result from household sleeping arrangements where older males and school-aged children may sleep separately from mothers with young children, as suggested in other studies [53, 54]. Another element of sleeping arrangements may concern the size of the home, as households with only one room had higher odds of malaria than households with two or more rooms. In one-room households, it is possible that bed nets cannot be hung properly in such confined spaces and occupants may touch the net while sleeping, allowing mosquitoes to bite through the mesh or not be properly covered while sleeping [55, 56].

Local environmental vegetation around the household increased the odds of malaria infection. This effect remained robust despite adjusting for the most important variable within our models, non-linearly modeled age, and other sociodemographic characteristics. Higher NDVI values represent greater vegetation density and may serve as a proxy indicator for a mosquito-suitable environment immediately around the home. This interpretation aligns with other studies in SSA reporting positive NDVI-malaria associations, though with varying magnitudes (OR 1.5– 4.0) depending on spatial resolution and context [10, 16, 17, 20, 57, 58]. The NDVI effect may also capture social structure, as households on the village periphery where vegetation is denser might represent poorer households or renters who cannot afford central locations [59–61]. Our community mapping analysis showed lower NDVI values in village centers where trading centers and main roads are located, while peripheral areas showed higher vegetation density. Landform type remained significant after adjusting for NDVI, suggesting that landform and NDVI capture different elements or locations of environmental exposure. NDVI quantifies vegetation within 32– 64 m of households, while water landform reflects broader ecological features. The importance of the immediate household environment aligns with studies demonstrating micro-environmental conditions within 50–100 m associate with increased risk [20, 62–65].

We found mixed results for socioeconomic determinants. For occupational exposure, fishermen were not more likely to have malaria despite higher exposure to vegetated shorelines and outdoor activities. The effect observed in unadjusted analyses was attenuated after age and gender adjustment, as fishermen were predominantly older males, who have likely developed gradual immunity to malaria [66, 67]. Further, most fishing occurs during daylight hours when bites come primarily from other mosquito species along vegetated shorelines, while the dominant malaria vectors *Anopheles funestus* and *An. gambiae* bite at night [68–71]. Night fishing, though coinciding with vector-biting periods, is mainly conducted in deep waters with limited vegetation and potentially fewer mosquitoes by fishermen wearing thicker clothing for warmth that provides bite protection [72].

Further, farmers had lower infection odds than those without occupations. Farmland ownership outside villages means farmers spend more time away from high-risk shoreline areas [73]. In contrast to other studies, we found no association of years of education or home quality [60, 74–76]. Importantly, home quality was no longer significant after adjusting for NDVI, which captured the household microenvironment, suggesting that future research should not study these two variables in isolation.

Individuals belonging to households in closer spatial proximity to government health centers were less likely to have malaria. This finding may represent access to malaria treatment and preventive interventions [77–79]. Importantly, healthcare access in our study communities for malaria operates through two main channels for older children and adults (excluding the integrated management of childhood illnesses for children aged *<* 5 years). Government health centers provide free antimalarial treatment covering the full treatment course (though stock-outs are common), and private drug shops sell antimalarials at market prices, which are often higher than recommended retail prices [80–82]. We found that only the distance to government health centers was relevant for malaria status, while proximity to private drug shops showed no association, potentially reflecting community preferences for free treatment despite greater distances, as other studies and field observations have highlighted [79, 83]. Further, this finding may also represent the use of authentic antimalarials at government facilities, non-expired medicines, and access to adequate full courses for malaria treatment [80, 81, 84].

The reduction in the odds of infection observed in individuals recruited in 2024 warrants careful interpretation. Bed net coverage in our study areas approached or exceeded the Uganda Malaria Reduction and Elimination Strategic Plan 2021–2025 target of 90% population coverage [31]. In Pakwach, which had the highest malaria burden, a mass LLIN distribution campaign occurred in October 2023 as part of Wave 3 of the national campaign led by the Ministry of Health with support from nongovernmental organizations [85, 86]. Coverage in Pakwach increased significantly from 84.6% to 96.0% following this campaign, while Buliisa and Mayuge maintained consistently high coverage (*>* 93%) throughout the study period.

Our work contributes methods for processing remote sensing data by combining environmental data with household-level infection data, and by validating spatial patterns against field observations [13, 14, 16, 17, 20]. We used high spatial resolution, open-access satellite reflectance data at the household scale, coupled with a smoothing and validation framework to contextualize NDVI values to the local area and reduce pixel artifacts. Satellite imagery is used widely for malaria modeling mostly at coarser resolutions (250 m*×*250 m to 5 km*×*5 km grids) using climate (e.g., Land Surface Temperature) and spectral indices (e.g, NDVI) for district- or country-level risk mapping [13, 14, 16, 17, 20, 87–90]. The data available from the Landsat and Sentinel missions, mapping at 10–30 m resolutions, capture heterogeneity relevant to local sub-district level programs [17, 21, 91]. However, when applying reflectance-based indices at sub-district or even sub-village level scales, pixel-level noise, cloud cover, mixed-pixel effects at water–land boundaries, and grid artifacts can degrade data quality [92, 93]. We addressed these challenges by using a hexagonal lattice and applying Gaussian neighborhood smoothing. Smoothing strengthened associations with infection relative to unsmoothed indices. We complemented the satellite data with targeted ground observations, comparing indices with field malacology and participatory community maps. These observations helped identify artifacts and confirm that higher vegetation aligns with non-beach landforms and locally identified features. Satellite data offer broad coverage, but benefit from scale-appropriate preprocessing and selective validation; without these, micro-epidemiological risk can be mischaracterized [17, 21, 94]. Our findings also intersect with efforts to build operational early warning and decision-support systems [21]. Our household-level framework complements these developments by connecting individual and household risk with environmental structure at 10–30 m scales. This methodology is readily scalable for national malaria programs as Sentinel-2 data are freely accessible globally through the Copernicus Data Space Ecosystem, with continuous acquisition every 5 days ensuring temporal coverage for ongoing surveillance and risk mapping [95, 96].

This study has several strengths. We combined individual, household, and village data across three districts, applied a modeling strategy that captured non-linearity in age effects, and implemented a pragmatic geospatial pipeline using open data that can be reproduced in low-resource settings. Limitations include the lakeside focus, which may limit generalizability to inland settings, the cross-sectional design that cannot distinguish prevalent from incident malaria cases, the exclusion of children *<* 5 years of age, and self-reported LLIN use. The malacology protocol excluded ephemeral habitats, potentially underestimating breeding availability during rainy seasons [15, 97, 98]. As with any spatial analysis, the Modifiable Areal Unit Problem remains a consideration, although consistent tessellation and sensitivity analyses mitigated its impact. Finally, reflectance indices vary with seasonality and cloud cover; our pre-processing and validation reduce, but do not eliminate, these sources of error.

## Conclusion

Understanding age-dependent malaria dynamics is important for optimizing control strategies in high-burden settings. We found that malaria prevalence peaks at 10 to 11 years of age in Ugandan lakeside communities, with a substantial burden of asymptomatic infections persisting into adulthood. Household-level vegetation density and distance to government health centers were associated with increased odds of infection. If validated elsewhere, our fine-scale environmental indices could guide community health worker deployment and resource allocation in high-risk areas within districts. Importantly, our findings support expanding existing malaria interventions or trials of new interventions beyond children under five to include school-based screening programs for pre-adolescents while maintaining adult surveillance to address the asymptomatic adult reservoir.

## Data Availability

The raw data are protected and are not available due to data privacy and ethics restrictions. All relevant metadata and supplementary material are provided within the manuscript. The satellite data used in this study are publicly available, free of charge, and can be accessed from the Copernicus Open Access Hub. The code to reproduce the pipeline is available as supplementary material.

## Declarations

## Ethics Approval

Data collection and use were reviewed and approved by Oxford Tropical Research Ethics Committee (OxTREC 509-21), Vector Control Division Research Ethics Committee of the Uganda Ministry of Health (VCDREC146), and Uganda National Council for Science and Technology (UNCST HS 1664ES).

## Consent for publication

## Author Contributions

Conceptualisation: MML GFC.

Data curation: MML, NBK, VT, PK, AN, AM, BN, and GFC.

Formal analysis: MML.

Funding acquisition: MML and GFC.

Investigation: MML.

Methodology: MML, CAD, and GFC.

Project administration: NBK and GFC.

Resources: NBK and GFC.

Software: GFC.

Supervision: CAD and GFC.

Validation: MML.

Visualisation: MML.

Writing–original draft: MML.

Writing–review & editing: MML, NBK, VT, PK, AN, AM, BN, CAD, and GFC.

## Acknowledgements

We are grateful for the engagement and participation of all study participants and local communities involved in this research. We extend our thanks to all field teams, including the malaria technicians, auxiliary workers, and community members who contributed to the development of community maps and data collection efforts. We thank the SchistoTrack group for valuable feedback and insights during group meetings and conversations, and particularly acknowledge Dr Melissa Iacovidou for her valuable input and discussion, which helped improve this analysis and manuscript.

## List of Abbreviations

CI: Confidence Interval
CV: Coefficient of Variation
EVI: Enhanced Vegetation Index
GAMM: Generalized Additive Mixed Model
ICC: Intra-class Correlation Coefficient
IQR: Interquartile Range
IRS: Indoor Residual Spraying
LLIN: Long-Lasting Insecticidal Net
LST: Land Surface Temperature
MAP: Maximum a Posteriori
MNDWI: Modified Normalized Difference Water Index
NDVI: Normalized Difference Vegetation Index
NDWI: Normalized Difference Water Index
NPV: Negative Predictive Value
ODK: Open Data Kit
OR: Odds Ratio
OxTREC: Oxford Tropical Research Ethics Committee
PPV: Positive Predictive Value
RDT: Rapid Diagnostic Test
REML: Restricted Maximum Likelihood
ROC: Receiver Operating Characteristic
SSA: Sub-Saharan Africa
UNCST: Uganda National Council for Science and Technology
UNICEF: United Nations International Children’s Emergency Fund
UTM: Universal Transverse Mercator
VCDREC: Vector Control Division Research Ethics Committee
WASH: Water, sanitation, and hygiene
WHO: World Health Organization

