## Supplementary material for "Non-linear age dynamics of malaria infection and fine-scale environmental exposure in rural Uganda"

#### Supplementary Information

##### S1 Text: Malaria Testing Procedures

Technicians read RDT results after a minimum of 15 minutes and no longer than 30 minutes. Tests were considered invalid if only the test line appeared without a control line or if the sample did not move across the entire test strip. Invalid tests were repeated until a valid result was obtained. A 10% random sample underwent quality control re-reading by a senior technician.

Blood film slides were read by one technician following WHO guidelines. Parasitaemia was calculated based on thick film counts according to the formula specified in WHO guidelines for malaria microscopy [1].

At clinical stations, trained government nurses from the participant's district assessed current symptoms and asked about fever episodes within the past month. Of 4308 participants who obtained malaria tests, 4219 (97.9%) additionally obtained malaria slide readings.

##### S2 Text: Covariate Definitions and Categorizations

Age was recorded in completed years at the time of recruitment. Gender was recorded as binary (male/female). Years of formal education ranged from 0 to 14, representing primary through secondary education levels. Occupation was based on the main income-earning activity, as individuals might have several economic activities. We recorded only the primary source of income to assess occupational risk factors.

For each individual, we first determined the majority tribe and religion within their specific village based on the full sample of participants from that village. Each participant was then coded as a binary variable indicating whether they belonged to their village's majority tribe (1 = yes, 0 = no) and similarly for religion. This approach allowed us to assess whether minority status within one's own community was associated with differential malaria risk.

Household social status was encoded as a binary variable at the household level, equal to one if any household member held a recognized local leadership position, and zero otherwise. Eligible positions included: Executive positions on the local council (Chairman, Vice-Chairman, Secretary), Specialized secretariat roles (Gender Secretary, Youth Council), Community leadership posts (Religious Leader, Clan Leader), Members of Village Health Team, and Beach Management Committee members.

The home quality score (ranging 3–12) was calculated as the sum of three components: Wall material (1 = Mud sticks, 2 = Plastic, 3 = Metal, 4 = Bricks or cement), Roof type (1 = Grass or Papyrus, 2 = Sticks, 3 = Plastic, 4 = Metal), Floor type (1 = Mud, 2 = Plastic, 3 = Wooden Planks, 4 = Bricks or cement).

All geographic coordinates were projected to Universal Transverse Mercator (UTM) zone EPSG:32636 for distance calculations. Where field-collected coordinates and OpenStreetMap facilities were located within 10 meters of each other, we prioritized the field-collected data.

Water body types were assigned based on the nearest malacology survey site to each household. Classifications included: Beach, Lake marsh, River/river marsh, Pond/swamp, and Rice paddy.

#### S3 Text: Environmental Data Processing

Environmental data processing followed a two-stage workflow: (1) satellite imagery acquisition and index calculation in Google Earth Engine, followed by (2) spatial change-of-support, smoothing, and extraction in R.

##### Stage 1: Google Earth Engine Processing

We used the Sentinel-2-L2A (`COPERNICUS/S2_SR_HARMONIZED`) image collection in Google Earth Engine. We first filtered the collection for each two-month analysis period to gather scenes with less than 20% cloud cover (`CLOUDY_PIXEL_PERCENTAGE`), then applied a pixel-level mask to each remaining image using the Sentinel-2 Scene Classification Layer (SCL), removing pixels classified as cloud shadow (class 3), medium probability cloud (class 8), and high probability cloud (class 9). To create a single cloud-free observation for each period, we generated a (best-available) median composite from all valid pixels. Finally, we calculated the indices and exported the data at a 10 m spatial resolution.

Four vegetation and water indices were computed directly in Google Earth Engine using the following formulas [2]:

$$\text{NDVI} = \frac{\text{NIR} - \text{Red}}{\text{NIR} + \text{Red}}, \tag{1}$$

$$\text{EVI} = 2.5 \cdot \frac{\text{NIR} - \text{Red}}{\text{NIR} + 6 \cdot \text{Red} - 7.5 \cdot \text{Blue} + 1}, \tag{2}$$

$$\text{NDWI} = \frac{\text{Green} - \text{NIR}}{\text{Green} + \text{NIR}}, \tag{3}$$

$$\text{MNDWI} = \frac{\text{Green} - \text{SWIR}_1}{\text{Green} + \text{SWIR}_1}. \tag{4}$$

##### Stage 2: R-based Spatial Analysis

We performed a change-of-support from the native square raster (10 m Sentinel-2) to hex cells using an areal-weighted mean with fractional overlap. Let $x_r$ denote the value in raster pixel $r$, $H_i$ denote hexagon $i$

with area $A_i$, and $A_{ir} = \text{area}(H_i \cap r)$ be the overlap area. The aggregated value is

$$\bar{x}_i = \sum_{r:\, H_i \cap r \neq \emptyset} w_{ir}\, x_r \quad \text{with} \quad w_{ir} = \frac{A_{ir}}{\sum_{r'} A_{ir'}} = \frac{A_{ir}}{A_i}. \tag{5}$$

This approach (i) handles partial coverage along water–land boundaries without hard pixel thresholds, (ii) dampens pixel-level speckle/cloud artifacts, and (iii) provides a common analysis support for linking environmental indices to households and malacology sites. Implementation used R `exactextractr` (`exact_extract` with coverage fractions as weights). The same procedure applies to any base raster (e.g., 30 m Landsat), ensuring method generality across sensors.

Aggregating to hex cells before smoothing stabilizes values at the intended analysis scale, reduces mixed-pixel effects at edges, and ensures that neighborhood operations act on comparable areal units rather than raw pixels.

We pre-computed spatial relationships on the hex lattice:

- **First-order neighbors** ($\mathcal{N}_i^{(1)}$): hexagons sharing an edge with $H_i$ (six neighbors);
- **Second-order neighbors** ($\mathcal{N}_i^{(2)}$): hexagons adjacent to first-order neighbors but not to $H_i$ (twelve neighbors).

Let $d$ denote the normalized centroid distance on the hex lattice: $d = 0$ (center), $d = 1$ (first ring), $d = \sqrt{3}$ (second ring).

We used Gaussian weights as a function of $d$:

$$w(d) \;=\; \exp\!\left(-\frac{d^2}{2}\right), \tag{6}$$

giving $w_0 = 1.0$ (center), $w_1 = \exp(-\frac{1}{2}) \approx 0.607$ (first ring), and $w_2 = \exp(-\frac{3}{2}) \approx 0.223$ (second ring).

For each hex $i$, the smoothed value was computed as

$$\tilde{x}_i \;=\; \frac{w_0 x_i \;+\; \sum_{j \in \mathcal{N}_i^{(1)}} w_1 x_j \;+\; \sum_{k \in \mathcal{N}_i^{(2)}} w_2 x_k}{w_0 \;+\; |\mathcal{N}_i^{(1)}|\, w_1 \;+\; |\mathcal{N}_i^{(2)}|\, w_2}. \tag{7}$$

We computed both raw (areal-weighted) and smoothed surfaces for each index and year (2022, 2023, 2024), and then linked values to household and malacology locations by recruitment year.

Previous research has identified the micro-environment around households as an important factor for malaria transmission and risk, but called for finer-scale environmental measurements to characterize these effects [3, 4]. Research demonstrated that household-level malaria risk extends beyond individual behaviors to include neighborhood effects [5, 6, 7], which also touches on findings from mosquito flight ranges that suggest there exist heterogeneous exposure risk within districts and villages [3, 8].

All rasters were reprojected to UTM (EPSG:32636) to enable appropriate distance and area calculations and to avoid angular distortion. We then generated a hexagonal analysis lattice covering each district. We used a pointy-topped orientation. For a target cell area $A \approx 900\,\text{m}^2$, the hex edge length is

$$a \;=\; \sqrt{\frac{2A}{3\sqrt{3}}} \;\approx\; 18.6\,\text{m}. \tag{8}$$

Under a pointy-topped layout, the point-to-point (vertical) span equals $2a \approx 37.2\,\text{m}$ and the flat-to-flat (horizontal) span equals $\sqrt{3}\,a \approx 32.2\,\text{m}$. The neighborhood topology (six first-order and twelve second-order neighbors) is unchanged by orientation.

#### S4 Text: Model Specification and Estimation

##### Data Collection and Management

Open Data Kit (ODK) Collect (versions 2022.4.2, 2023.2.4, and 2024.1) was used for data collection on Android devices (software version Android 9 and 10) and ODK Central (versions 2022.3.1, 2023.2.1, and 2024.2.1) to manage data and perform quality control.

##### Statistical Testing

For comparisons between groups, we used Pearson's chi-squared tests for categorical variables and Wilcoxon rank-sum tests for continuous variables. Spearman's rank correlation was used for all correlation analyses, with rank-biserial correlation calculated for associations between binary and continuous variables. All statistical tests were two-sided with significance set at $\alpha = 0.05$.

##### Spatial Analysis

To assess spatial clustering of malaria cases, we aggregated data to the household level, classifying households as positive if either sampled individual tested positive. We calculated Moran's $I$ statistic [9] across a range of $k$-nearest neighbor specifications ($k = 2$ to 50) with row-standardized spatial weights, implemented using the `spdep` package [10]. This range was selected to capture neighborhood structures within villages, which averaged approximately 42 unique households.

##### Variable Selection and Model Formulation

Prior to modeling, we examined pairwise Spearman's correlations among all candidate predictors, excluding variables with significant coefficients where $|r| > 0.6$. Based on these correlations and our validation analyses, we selected NDVI as the primary environmental predictor, excluding the other indices to avoid multicollinearity.

We utilized GAMMs to characterize the non-linear patterns of malaria RDT positivity over age [11]. Our model takes the form:

$$\eta_i = \beta_0 + \mathbf{X}_i\boldsymbol{\beta} + f_{\text{adapt}}(\text{age}_i) + f(\text{village}_{k(i)}) \tag{9}$$

where $\eta_i$ is the linear predictor for individual $i$ linked to the binary infection status through the logistic link function $g(\eta) = \log\left(\frac{\pi}{1-\pi}\right)$, $\mathbf{X}_i$ contains covariate values, $f_{\text{adapt}}$ is an adaptive smooth function for age, and $f(\text{village}_{k(i)})$ represents the random effect for the village containing individual $i$.

The response variable for each individual, $Y_i$, is assumed to follow an independent Bernoulli distribution, conditional on the probability of success $p_i$:

$$Y_i \mid p_i \sim \text{Bernoulli}(p_i). \tag{10}$$

The relationship between the expected value of the response, $p_i$, and the linear predictor, $\eta_i$, is defined by the logit link function:

$$\text{logit}(p_i) = \log\left(\frac{p_i}{1-p_i}\right) = \eta_i. \tag{11}$$

The linear predictor $\eta_i$ is a sum of fixed effects (parametric terms), a smooth function, and a random effect for the village:

$$\eta_i = \beta_0 + \sum_{j=1}^{P} \beta_j x_{ij} + f(\text{age}_i) + b_{k(i)} \tag{12}$$

where $\beta_0, \ldots, \beta_P$ are the fixed effect coefficients, $f(\text{age}_i)$ is the smooth function of age, and $b_{k(i)}$ is the random intercept for the village $k$ of individual $i$.

Model parameters were estimated by optimizing a penalized version of the log-likelihood via REML:

$$\hat{\boldsymbol{\beta}} = \arg\min_{\boldsymbol{\beta}} \left( -l(\boldsymbol{\beta}) + \frac{1}{2} \sum_j \lambda_j \boldsymbol{\beta}^\top S_j \boldsymbol{\beta} \right). \tag{13}$$

The penalty terms, $\frac{1}{2}\sum_j \lambda_j \boldsymbol{\beta}^\top S_j \boldsymbol{\beta}$, control the smoothness of the spline functions.

This modeling approach can be viewed from both frequentist and Bayesian perspectives [11, 12, 13, 14]. From a frequentist standpoint, we are minimizing a penalized log-likelihood; from a Bayesian viewpoint, this is equivalent to finding the maximum a posteriori (MAP) estimate for $\boldsymbol{\beta}$ under a Gaussian prior distribution, $\boldsymbol{\beta} \sim N(0, S_\lambda^-)$, where $S_\lambda = \sum_j \lambda_j S_j$ and $S_\lambda^-$ is a suitable pseudo-inverse (e.g., the Moore-Penrose generalized inverse) of the singular matrix $S_\lambda$ [11, 12, 15, 14].

In contrast to assigning $\lambda$ a hyperprior distribution as in the fully Bayesian approach, we estimate the smoothing parameter from the data, making this an empirical Bayes approach:

$$\text{Posterior for } \boldsymbol{\beta} \propto \text{Likelihood} \times \text{Prior distribution for } \boldsymbol{\beta} \tag{14}$$

$$p(\boldsymbol{\beta} \mid \mathbf{y}, \hat{\lambda}_{\text{age}}, \hat{\sigma}^2_{\text{village}}) \propto p(\mathbf{y} \mid \boldsymbol{\beta}) \times p(\boldsymbol{\beta}_{\text{fixed}}) \times p(\boldsymbol{\beta}_{\text{age}} \mid \hat{\lambda}_{\text{age}}) \times p(\mathbf{b} \mid \hat{\sigma}^2_{\text{village}}). \tag{15}$$

Prior distributions (conditional on estimated hyperparameters):

$$\boldsymbol{\beta}_{\text{fixed}} \sim N(\mathbf{0}, \boldsymbol{\Sigma}_{\text{fixed}}) \quad \text{(often diffuse or improper)} \tag{16}$$

$$\boldsymbol{\beta}_{\text{age}} \mid \hat{\lambda}_{\text{age}} \sim N(\mathbf{0}, (\hat{\lambda}_{\text{age}} \mathbf{S}_{\text{age}})^-) \tag{17}$$

$$b_{\text{village},k} \mid \hat{\sigma}^2_{\text{village}} \sim N(0, \hat{\sigma}^2_{\text{village}}). \tag{18}$$

Hyperparameter Estimation:

$$(\hat{\lambda}_{\text{age}}, \hat{\sigma}^2_{\text{village}}) = \underset{\lambda_{\text{age}}, \sigma^2_{\text{village}}}{\text{argmax}} \int p(\mathbf{y} \mid \boldsymbol{\beta}) p(\boldsymbol{\beta} \mid \lambda_{\text{age}}, \sigma^2_{\text{village}}) \, d\boldsymbol{\beta}. \tag{19}$$

The age effect was modeled using an adaptive smoothing spline with an order three penalty basis ($m = 3$), meaning that the smoothing parameter $\lambda$(age) is not constant but is itself represented as a smooth function using a spline with three basis functions, thereby allowing the degree of smoothness to vary flexibly over age. We set the basis dimension to $k = 20$ and confirmed adequacy using the `k.check()` diagnostic function. The selection of $m = 3$ was informed by exploratory analysis of the age-infection relationship through non-parametric visualizations and examination of smoothing parameter estimates across models with varying penalty basis dimensions ($m = 1$ through $m = 10$). We evaluated the smoothing parameter values $\lambda$ and visualized the penalty structure ($S \times \lambda$) using heatmaps [11, 16].

The adaptive smooth for age can be expressed as:

$$f_{adapt}(\text{age}_i) = \sum_{l=1}^{k} \gamma_l B_l(\text{age}_i) \tag{20}$$

where $B_l(\text{age}_i)$ are B-spline basis functions with dimension $k = 20$. The key feature of adaptive smoothing is that the penalty varies with $x$ (in our case age) through a basis expansion with dimension $m$:

$$\mathcal{P}_{adapt} = \sum_{j=1}^{m} \lambda_j \boldsymbol{\beta}^\top \mathbf{S}_j \boldsymbol{\beta} \tag{21}$$

where:

- $m = 3$ is the number of smoothing parameters (and the dimension of the penalty basis);
- $\lambda_1, \lambda_2, \lambda_3$ are the three smoothing parameters to be estimated;
- each $\mathbf{S}_j$ corresponds to one of the $m$ basis functions used to represent the spatially varying penalty.

The penalty matrices are constructed as $\mathbf{S}_j = \mathbf{D}^\top \text{diag}(\mathbf{B}_{\cdot j})\mathbf{D}$, where $\mathbf{B}$ is now an $m$-column matrix of B-spline basis functions that represents how the penalty varies across age, $\mathbf{B}_{\cdot j}$ is its $j$-th column (for $j = 1, \ldots, m$).

The village effects $f(\text{village}_k) = b_k$ are treated as random effects with $b_k \sim N(0, \tau^2)$, where the penalty parameter $\lambda = \sigma^2/\tau^2$ controls the degree of shrinkage. For the age effect, the smoothing parameter controls the "wiggliness"/"smoothness" of the spline [11].

##### Posterior Distribution and Inference

We leverage the Bayesian interpretation for probabilistic interpretation of the smooth coefficients. Note that this probabilistic interpretation applies specifically to the penalized (smooth) components of the model, not to the unpenalized parametric coefficients. As shown in [11, 17, 12] using a large sample approximation, the posterior distribution of the parameters, conditional on the data $y$ and the estimated smoothing parameters $\hat{\lambda}$, can be expressed as:

$$\boldsymbol{\beta} | y, \hat{\lambda} \sim N(\hat{\boldsymbol{\beta}}, \boldsymbol{V}_\beta). \tag{22}$$

The derivation follows from the penalized log-likelihood, denoted here as $l_p(\boldsymbol{\beta})$:

$$l_p(\boldsymbol{\beta}) = l(\boldsymbol{\beta}) - \frac{1}{2} \sum_j \lambda_j \boldsymbol{\beta}^\top S_j \boldsymbol{\beta} \tag{23}$$

where $l(\boldsymbol{\beta})$ is the standard log-likelihood.

The prior distribution is $\boldsymbol{\beta} \sim N(0, S_\lambda^-)$, which has density:

$$f(\boldsymbol{\beta}) \propto \exp\left\{-\frac{1}{2}\boldsymbol{\beta}^\top S_\lambda \boldsymbol{\beta}\right\}. \tag{24}$$

The posterior is proportional to likelihood times prior distribution:

$$f(\boldsymbol{\beta}|y) \propto f(y|\boldsymbol{\beta}) f(\boldsymbol{\beta}) = \exp\{l(\boldsymbol{\beta})\} \exp\left\{-\frac{1}{2}\boldsymbol{\beta}^\top S_\lambda \boldsymbol{\beta}\right\}. \tag{25}$$

Taking logarithms:

$$\log f(\boldsymbol{\beta}|y) = l(\boldsymbol{\beta}) - \frac{1}{2}\boldsymbol{\beta}^\top S_\lambda \boldsymbol{\beta} + C \tag{26}$$

where $C$ is the logarithm of the normalizing constant, which does not depend on the parameter $\boldsymbol{\beta}$ and ensures the posterior distribution integrates to 1.

To account for the uncertainty introduced by estimating the smoothing parameters, we used a corrected posterior covariance matrix ($V_{corrected}$) that accounts for the estimated $\hat{\lambda}$ using a first-order Taylor approximation [17].

We implemented the following algorithm proposed by [11] using components from the `mgcv` R package. For each of the $B = 2000$ posterior draws, indexed by $b$, we performed the following steps:

a. Draw $\beta_{\text{smooth}}$ from $\boldsymbol{\beta}_{\text{smooth}} \mid \boldsymbol{y}, \boldsymbol{\lambda} \overset{a}{\sim} N\left(\widehat{\boldsymbol{\beta}_{\text{smooth}}}, \boldsymbol{V}_{\text{corrected}}\right)$ using a random-walk Metropolis-Hastings sampler (with 1000 burn-in samples) or approximated samples from Gaussian Multivariate Normal.

b. Calculate predicted infection probability for each age value of interest: $\hat{p}_j^* = g^{-1}(\mathbf{X}\beta_j)$.

c. Store $\hat{p}_j^*$, a vector of predicted probabilities across ages.

For each age value, we can calculate the posterior mean, and 95% credible intervals using either the 2.5% and 97.5% percentiles across the posterior simulations ($\hat{p}_j^*$) or using the Gaussian approximation with the posterior mean and standard errors derived from the corrected covariance matrix, $\mathbf{V}_{\text{corrected}}$ [18, 19]. The obtained credible intervals, if averaged over the entire function, have frequentist coverage guarantees; however they may vary locally, especially in regions of high curvature where the choice of smoothing parameter is most important [19, 20].

#### S5 Text: Community Mapping Protocol

This protocol was used with the village chairman as the primary cartographer. The process was participatory, with input encouraged from other community members present.

1. **Locate trading center:**
    - Ask to be taken to the trading center of the village.
    - Take the GPS coordinates of the trading center using OSM or a GPS device.
2. **Introduction to the village chairman:**
    - Meet with the village chairman at the trading center.
    - Briefly explain the purpose of the mapping exercise:
        - "We are mapping the village to understand areas where malaria and schistosomiasis are spreading and how we can improve healthcare for everyone."
    - Emphasise the importance of their role and knowledge in creating an accurate map.
3. **Map orientation and division:**
    - Divide the map into 4 quadrants:
        - Top Left, Top Right, Bottom Left, Bottom Right.
    - Mark the trading center as the center point of the map.
    - Draw a North arrow and explain the orientation:
        - Use a GPS device to establish which direction is North.
        - Say, "We are facing North. This means that in front of us is North, behind us is South, to the left is West, and to the right is East."

- Explain North in a way the locals relate to (e.g., sunrise/sunset or landmarks if needed).

4. **Drawing instructions:**
   - Ask the chairman to draw items based on their location relative to the trading center:
     - If the item is in front and to the left, draw it in the top left quadrant.
     - If the item is in front and to the right, draw it in the top right quadrant.
     - If the item is behind and to the left, draw it in the bottom left quadrant.
     - If the item is behind and to the right, draw it in the bottom right quadrant.
     - If the item is directly to the left, right, in front, or behind, draw it on the dividing line between quadrants.

5. **Use of symbols:**
   - Provide a list of symbols and explain their meaning (e.g., 'X' for water contact points).
   - Ensure the chairman uses these symbols consistently for each feature.

6. **Adding key features:**
   - **Lake and river:**
     - "Show me where Lake Albert or River Nile is. Draw it on the map like this (point to symbol). How far is it from here to the water? Let's write the distance inside the drawing. Put an 'X' where people go to fetch water or swim."

   Repeat for following:
   - **Village roads (including dirt roads):**
     - Draw all the main roads, including paths used regularly by villagers.
     - "Can you draw the main roads? Show the paths people use often, even if they are dirt roads."
   - **Chairman's home:**
     - Mark the chairman's home and label it clearly.
     - "Please show me where the home of the chairman is."
   - **Primary school:**
     - Draw and label the location of the primary school.
     - "Where is the primary school (if there is any)?"
   - **Health center:**
     - Mark the health center's position
   - **Drug shop:**
     - Draw and label the drug shop's location.
   - **Church:**
     - Draw and label the church's location.
   - **Taps:**
     - Mark the locations of taps used by the community for water access.
   - **Public latrines:**
     - Indicate the position of public latrines on the map.
   - **Homes:**
     - Identify and label homes based on the following categories:
       - **Lived in the village their whole life:**
         - Mark and label these homes with point to symbol.
       - **Lived in the village for less than 5 years:**

- Mark these homes with point to symbol.
    - **Renters:**
      - Use another unique symbol to identify rented homes.
      - **Explanation:** Renters may face different challenges related to housing and sanitation compared to homeowners.

- **Seasonal features:**
  - **Flooding areas:**
    - Mark areas prone to flooding during the rainy season with a wavy line.
    - Ask the chairman to estimate the size of these areas and note their locations relative to key landmarks like homes, roads, or schools.
  - **Dry season paths:**
    - Draw paths used exclusively during the dry season (e.g., shortcuts, temporary routes).
    - Use a dashed line to differentiate these from permanent roads.
- **Standing water sites:**
  - Mark the location of any standing water bodies (e.g., ponds, ditches) and note the distance from the trading center.
  - **Explanation:** Standing water can be breeding grounds for mosquitoes and other vectors. Mapping these helps in understanding disease transmission risks.

7. **Verification:**

- Review the map with the chairman to confirm accuracy.
- Clarify any ambiguous placements or features.

### Supplementary Figures

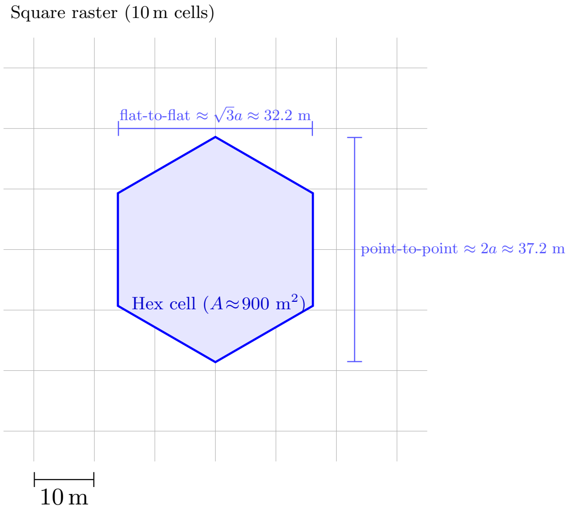


Figure S1: Areal-weighted change-of-support with a pointy-topped hexagon over 10 m pixels. Pixel contributions are weighted by fractional overlap $A_{ir}/A_i$.

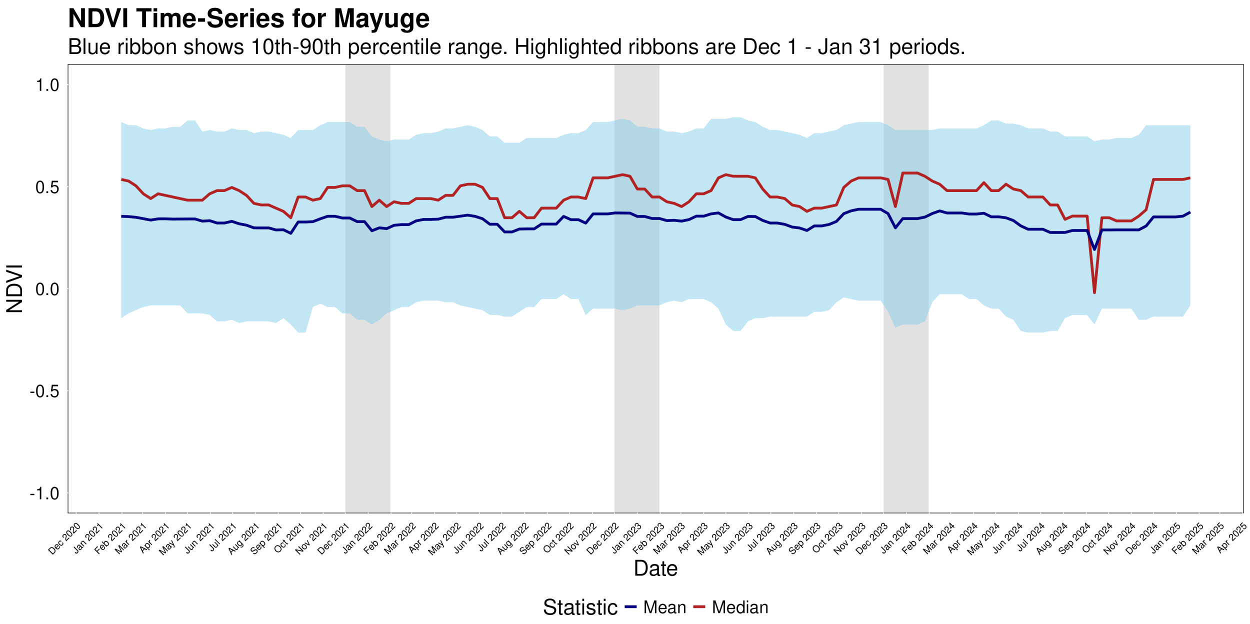


Figure S2: NDVI time-series for Mayuge district (2021–2025), generated using a 60-day moving window composite. The highlighted ribbons show the Dec 1 – Jan 31 analytical periods used in the main analysis, corresponding to the seasonal vegetation minimum.

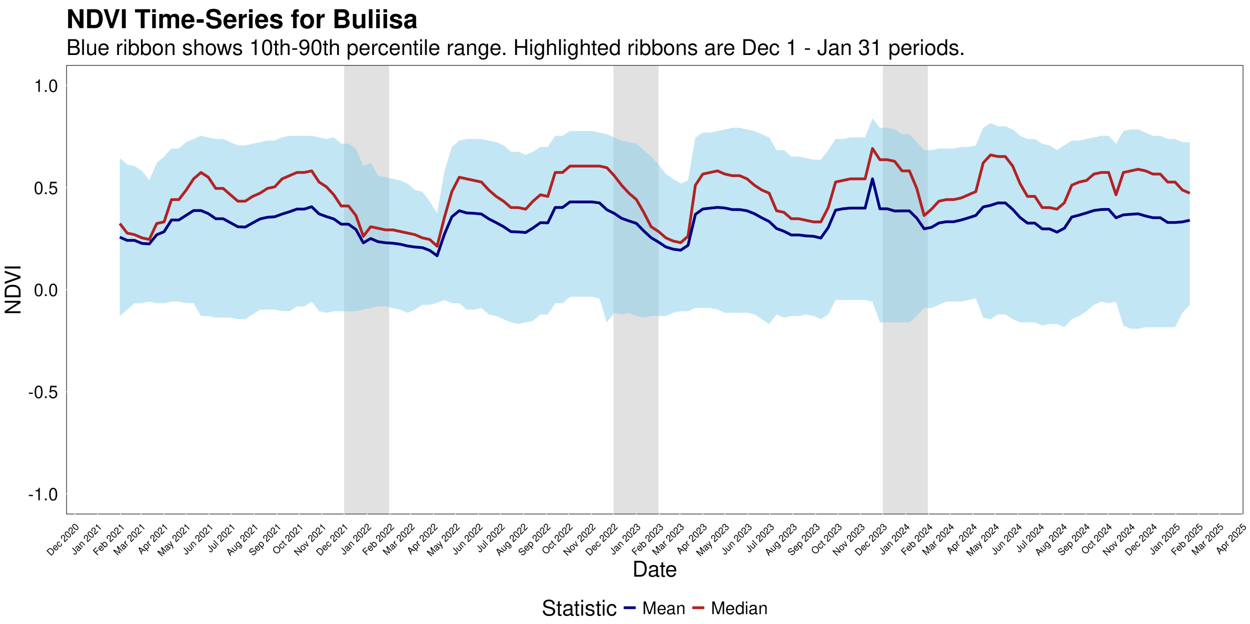


Figure S3: NDVI time-series for Buliisa district (2021–2025), generated using a 60-day moving window composite. The highlighted ribbons show the Dec 1 – Jan 31 analytical periods used in the main analysis, corresponding to the seasonal vegetation minimum.

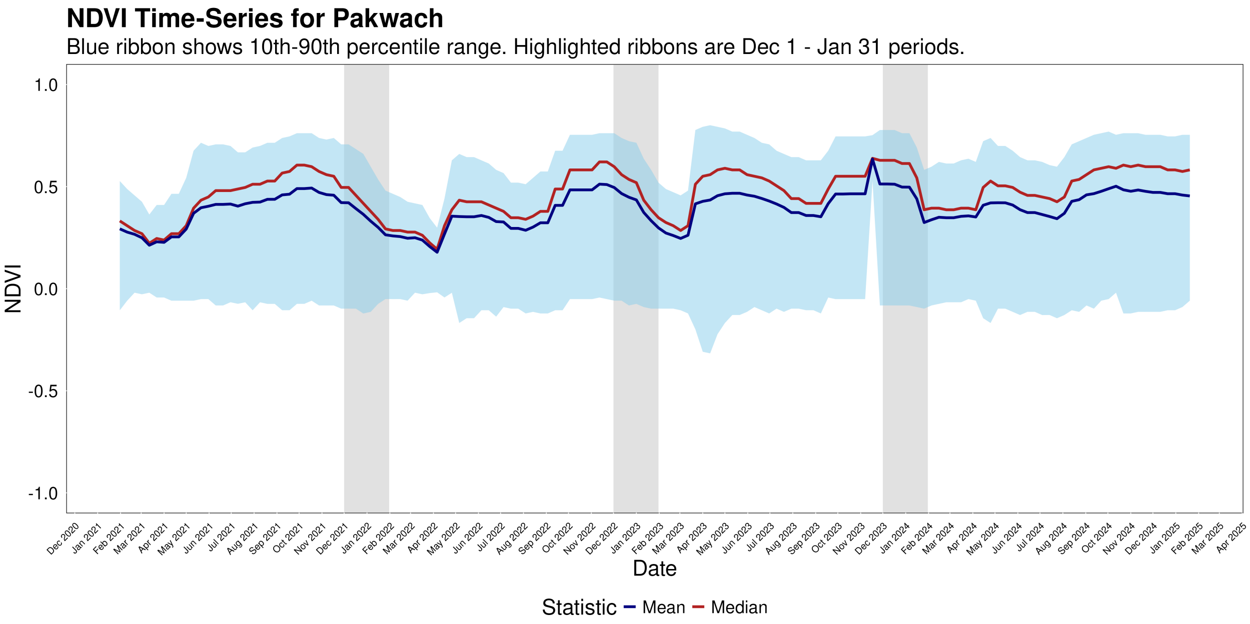


Figure S4: NDVI time-series for Pakwach district (2021–2025), generated using a 60-day moving window composite. The highlighted ribbons show the Dec 1 – Jan 31 analytical periods used in the main analysis, corresponding to the seasonal vegetation minimum.

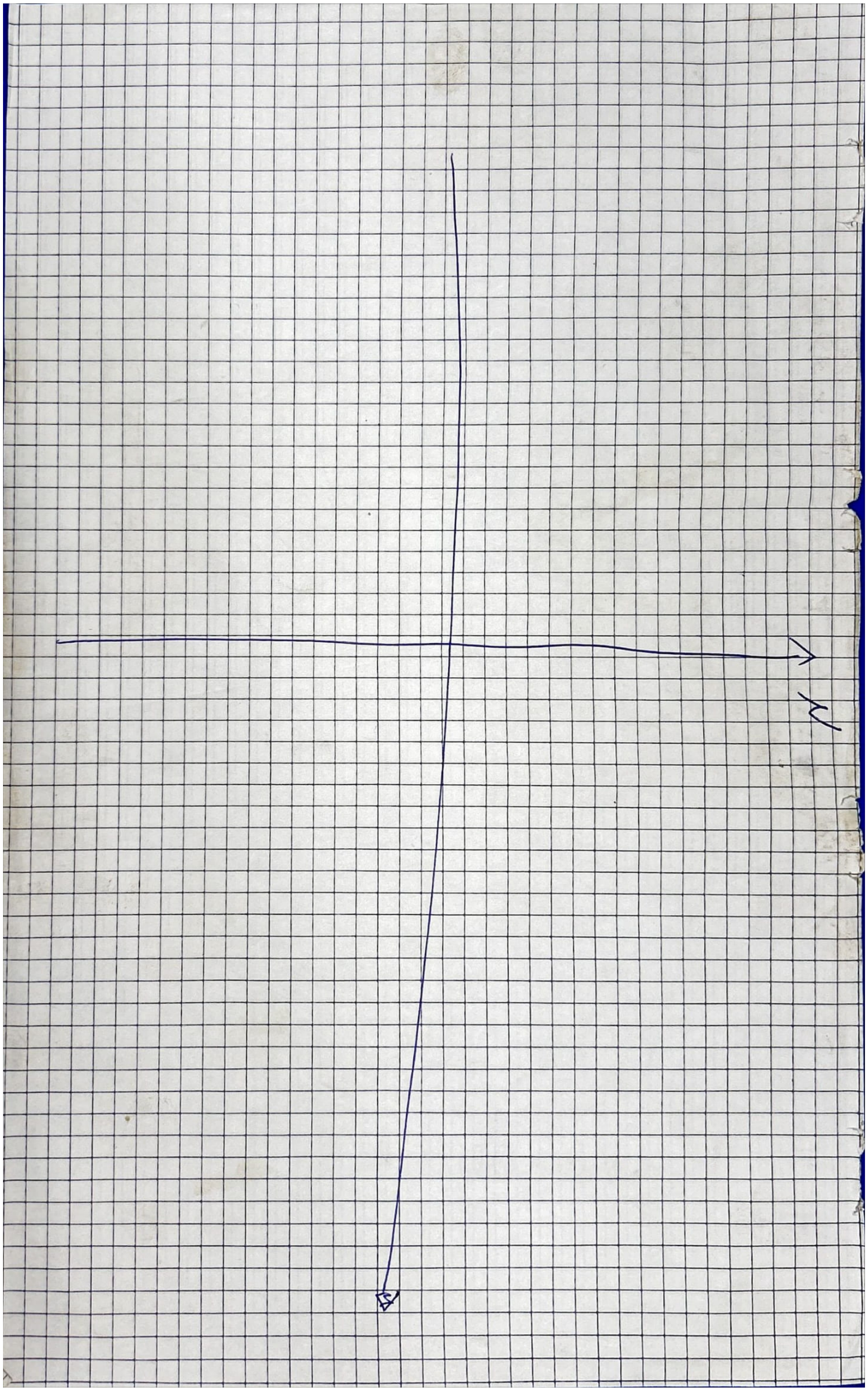

Figure S5: Community Mapping empty Grid.

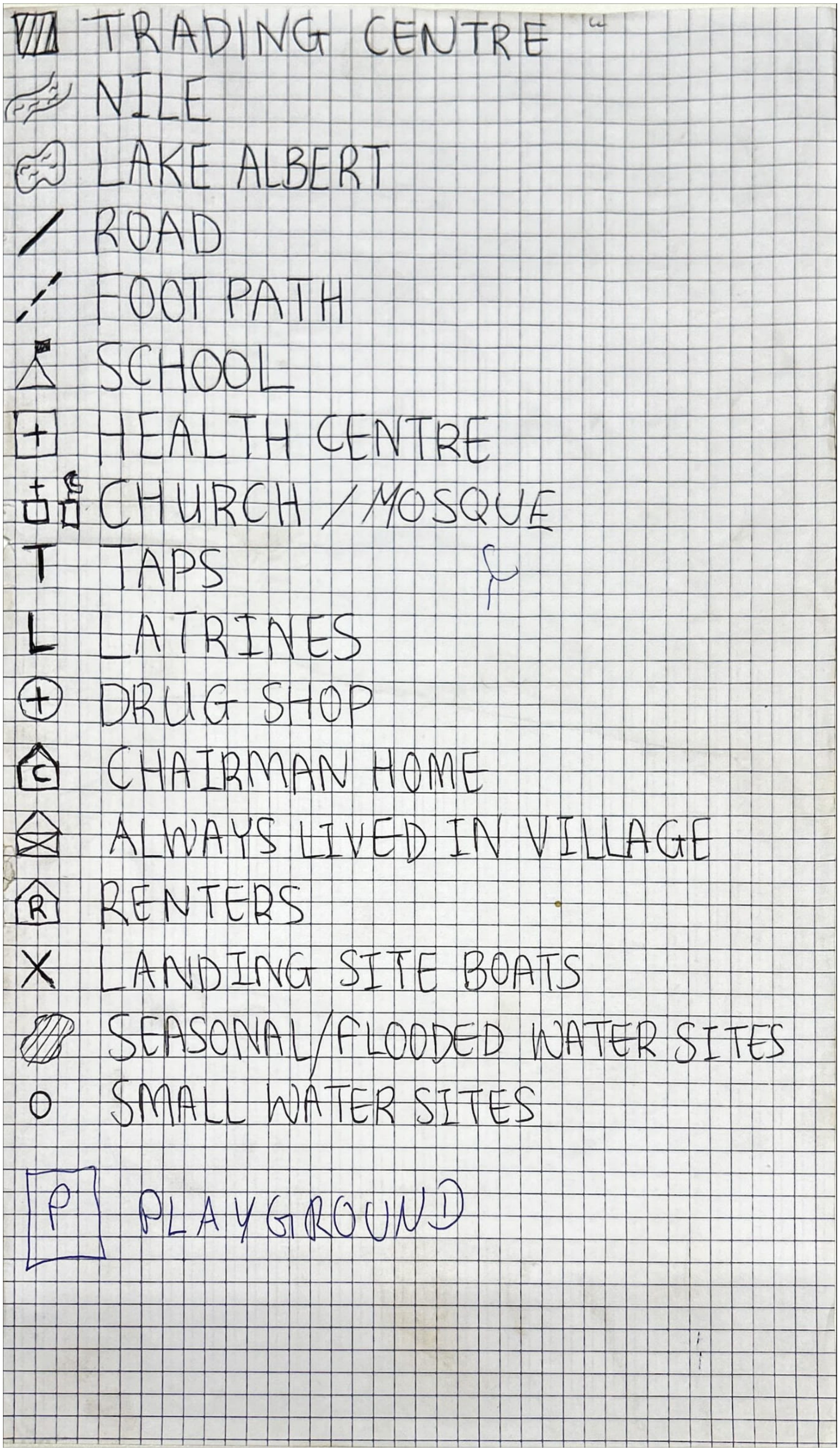


Figure S6: Community Mapping Symbol Legend.

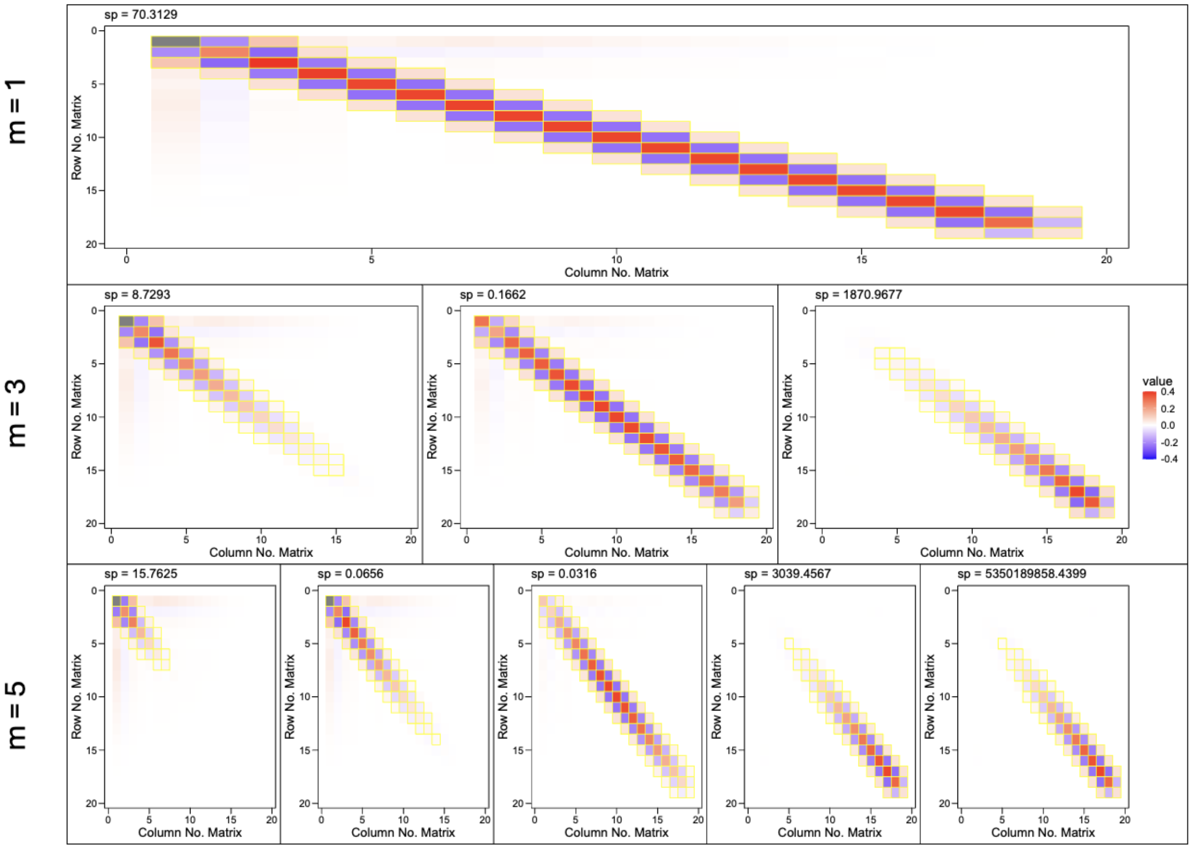


Figure S7: Structure of the adaptive penalty matrices for the age spline across different penalty basis dimensions ($m = 1, 3, 5$). Each heatmap visualizes a component penalty matrix ($\mathbf{S}_j$) scaled by its corresponding estimated smoothing parameter ($\hat{\lambda}_j$, labeled 'sp'). The final model utilized $m = 3$ (middle row), where the overall penalty is the sum of three such weighted matrices. This allows the wiggliness of the age-response curve to vary flexibly with age, in contrast to a standard smooth with a single penalty ($m = 1$, top row).

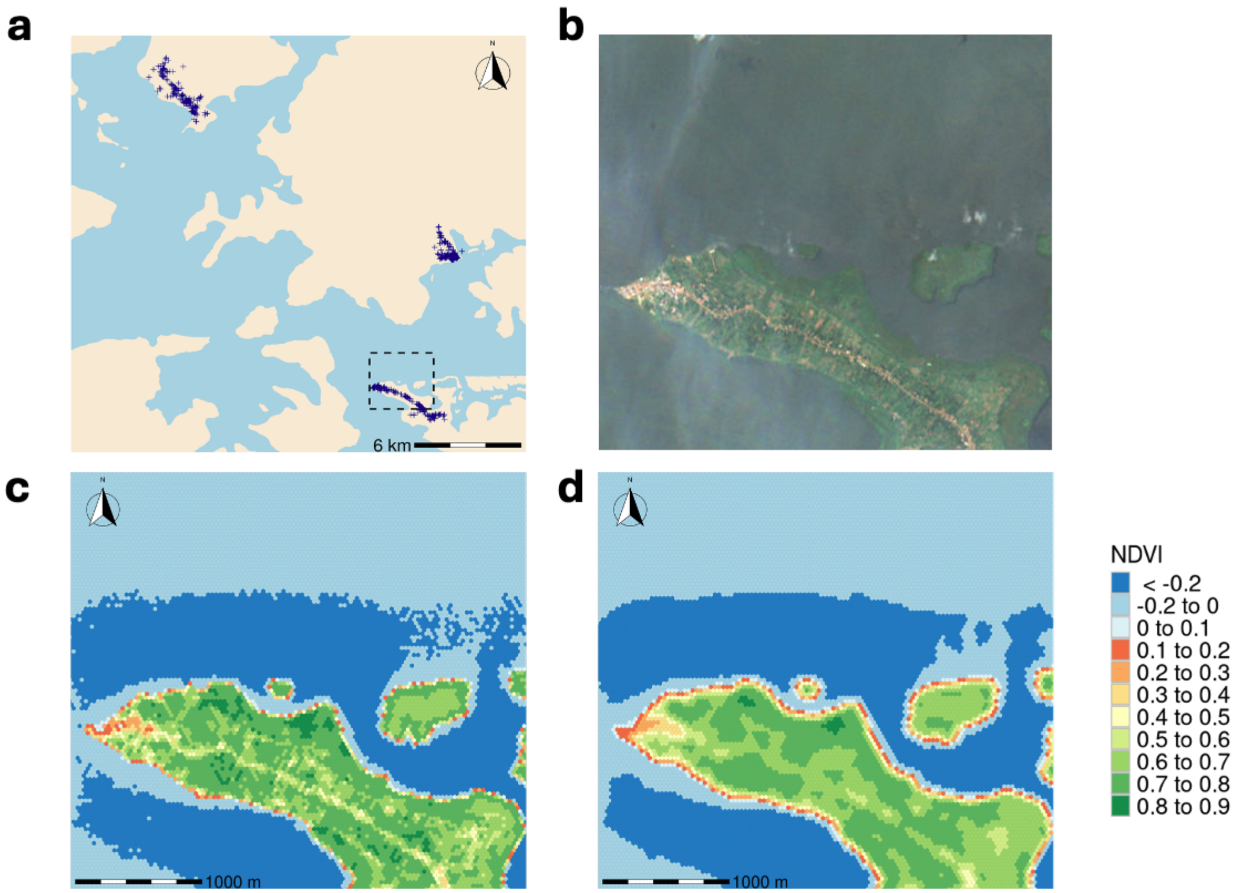


Figure S8: Spatial distribution of study sites and NDVI analysis. **(a)** Regional map showing household locations (blue markers). **(b)** Sentinel-2 satellite imagery of the area outlined in (a). **(c)** Unsmoothed NDVI values. **(d)** Spatially smoothed NDVI used for statistical analysis. The color gradient in (c) and (d) indicates vegetation density, from low (red) to high (green).

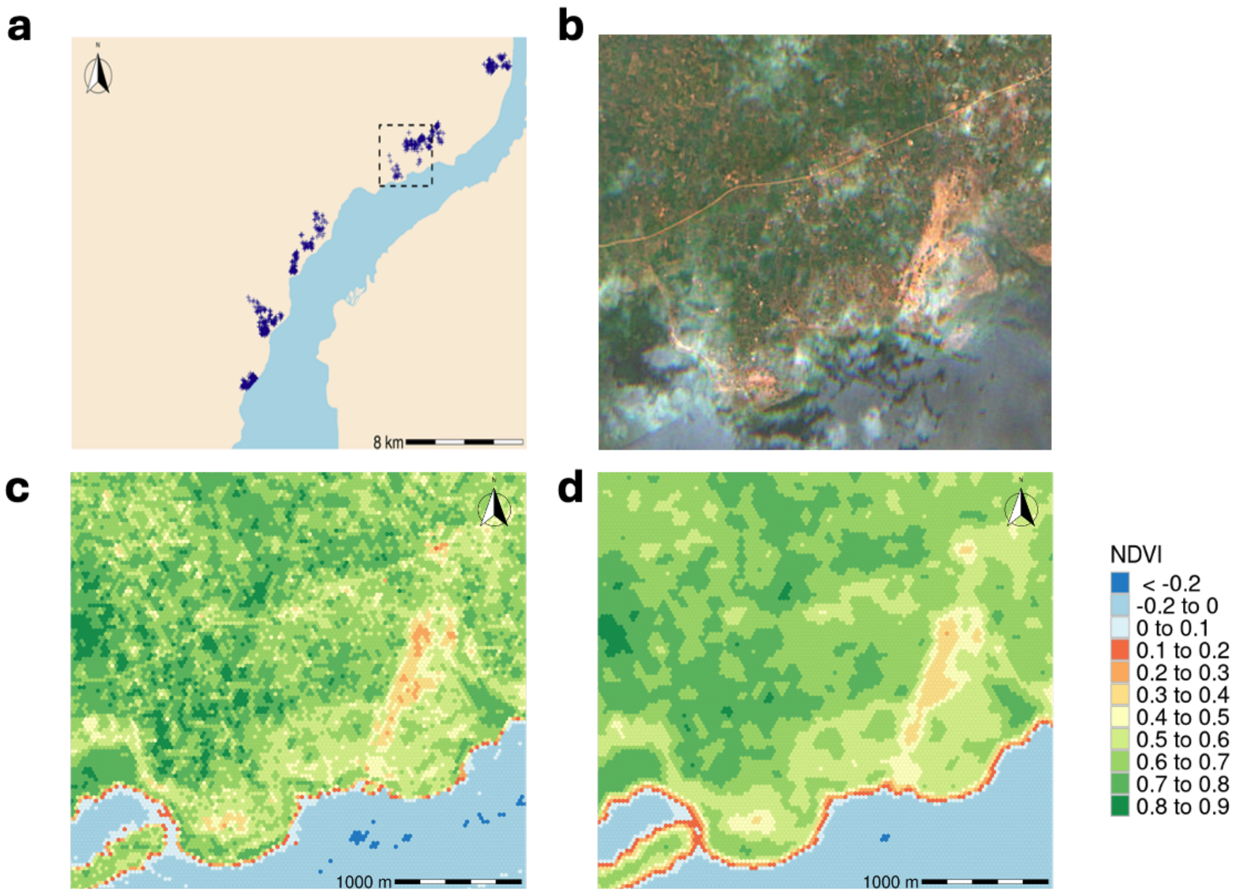


Figure S9: Spatial distribution of study sites and NDVI analysis. **(a)** Regional map showing household locations (blue markers). **(b)** Sentinel-2 satellite imagery of the area outlined in (a). **(c)** Unsmoothed NDVI values. **(d)** Spatially smoothed NDVI used for statistical analysis. The color gradient in (c) and (d) indicates vegetation density, from low (red) to high (green).

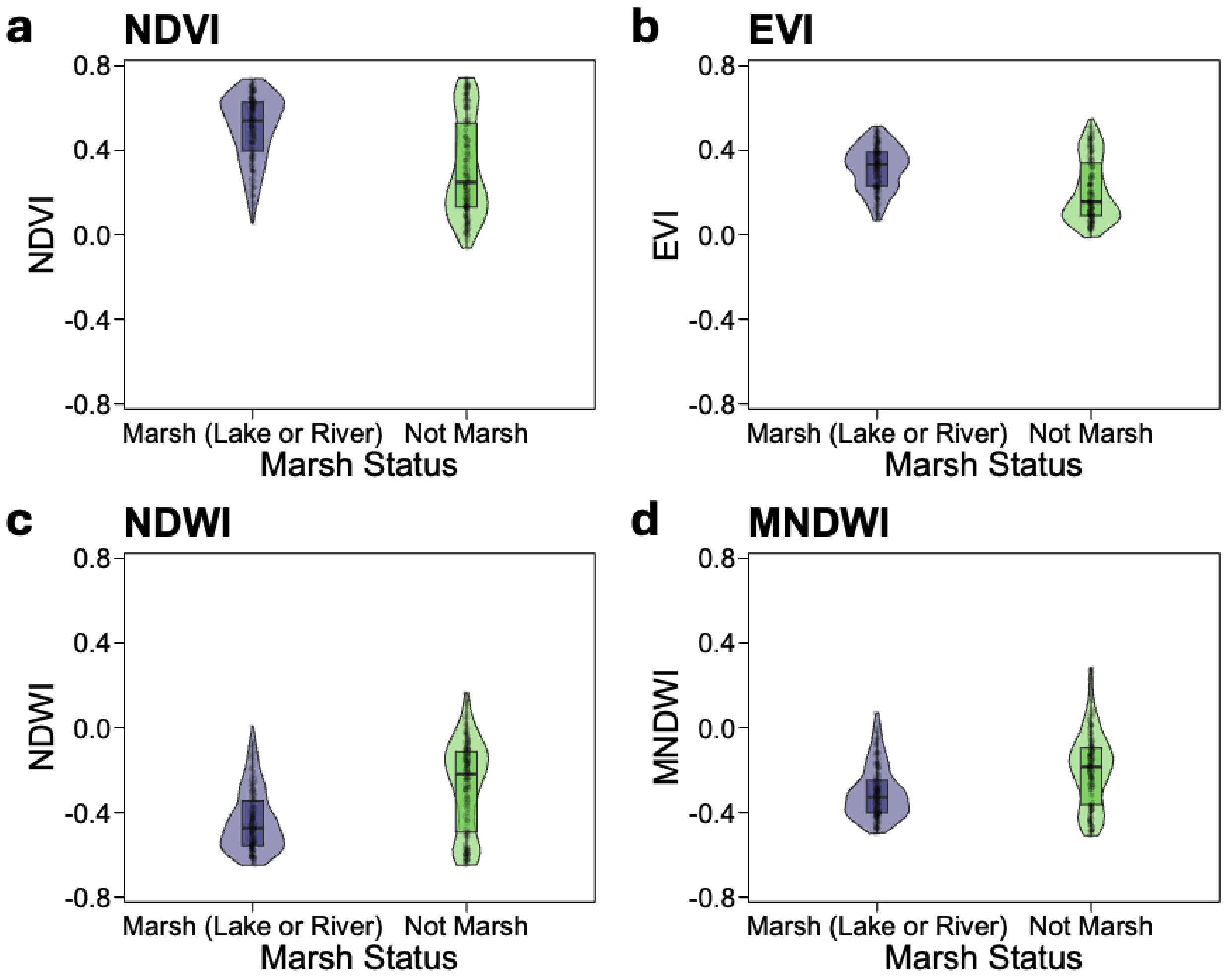


Figure S10: Validation of environmental indices against ground truth ecological data for malacological survey sites. The figure uses violin plots to show the distribution of each index. Each "violin" combines a box plot (inner white dot and black bar) with a kernel density plot (the outer shape), illustrating the median, interquartile range, and probability density of the data. The plots compare values for (a) NDVI, (b) NDWI, (c) MNDWI, and (d) EVI between marsh and non-marsh sites.

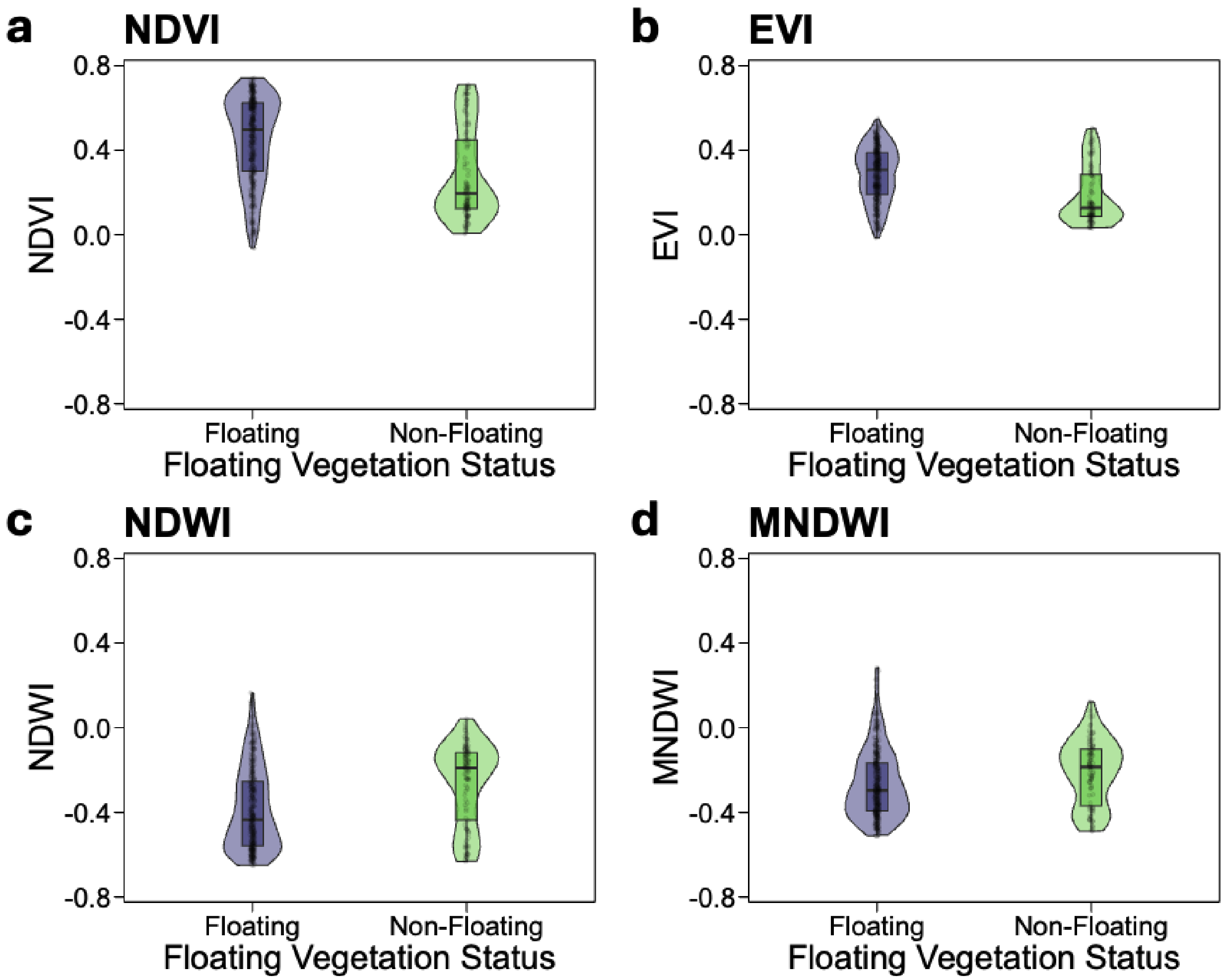


Figure S11: Validation of environmental indices against ground truth ecological data for malacological survey sites. (a) NDVI by floating vegetation versus non-floating vegetation status. (b) NDWI by floating vegetation versus non-floating vegetation status. (c) MNDWI by floating vegetation versus non-floating vegetation status. (d) EVI by floating vegetation versus non-floating vegetation status.

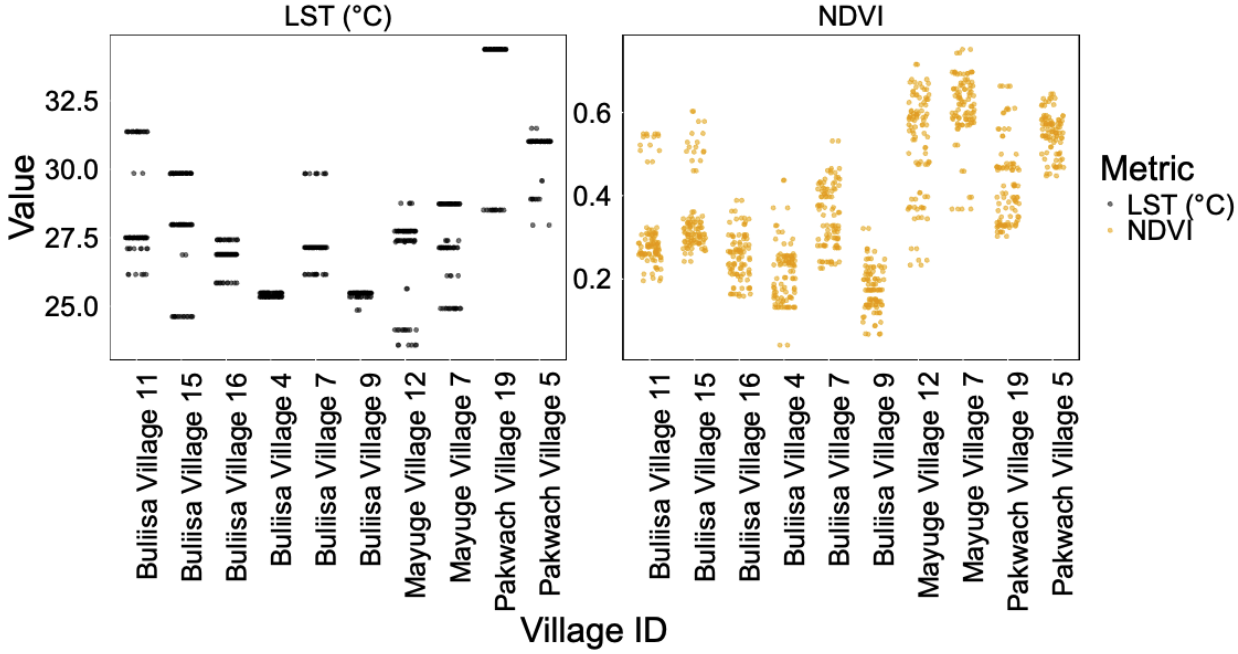


Figure S12: Distribution of Land Surface Temperature (LST) and Normalized Difference Vegetation Index (NDVI) values across study villages. Each point represents a household location. The panels illustrate the much coarser spatial resolution of the MODIS LST data (1000 m, left panel) compared to the Sentinel-2 NDVI data (10 m, right panel).

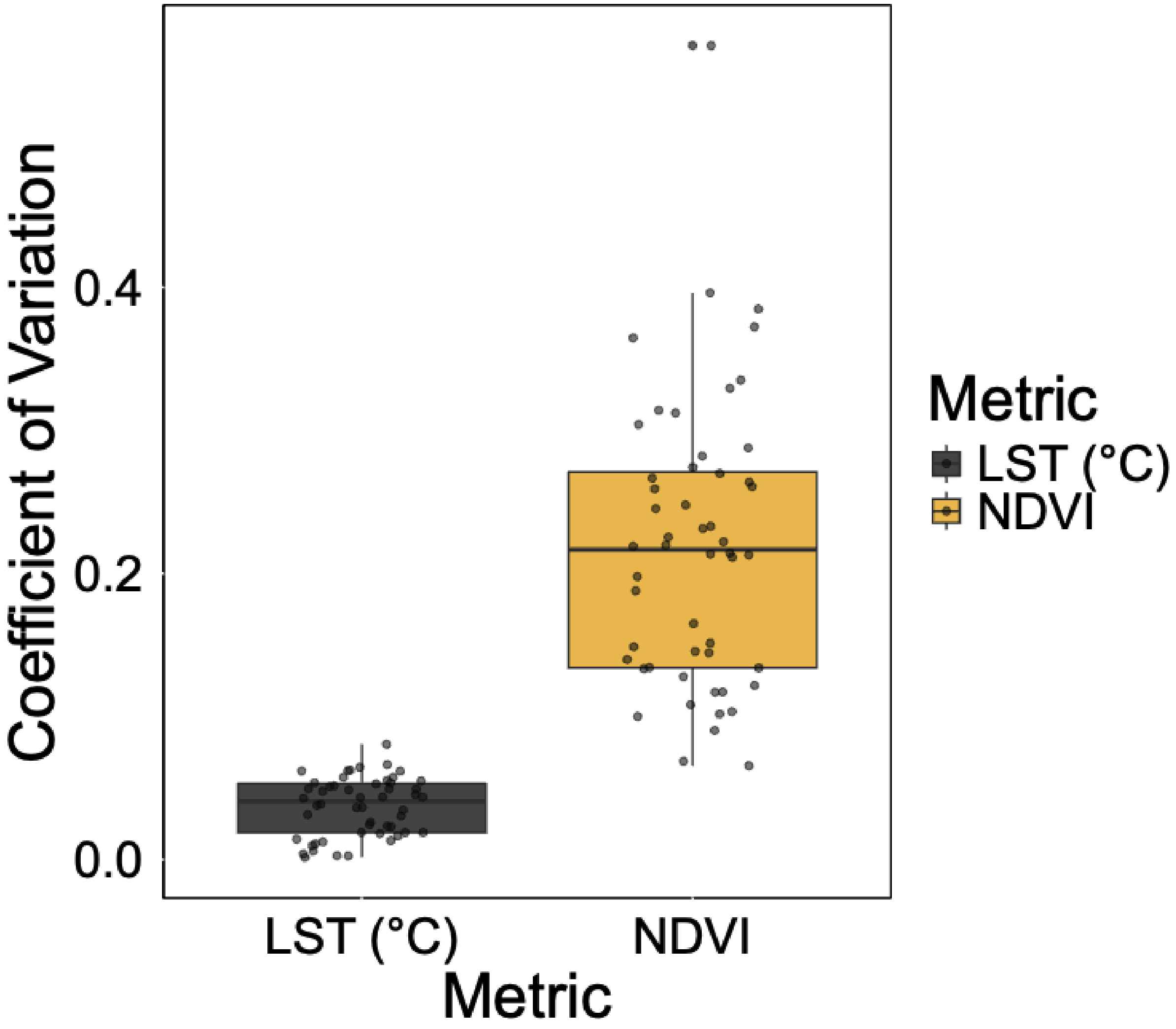


Figure S13: Comparison of the within-village coefficient of variation (CV) for Land Surface Temperature (LST) and NDVI.

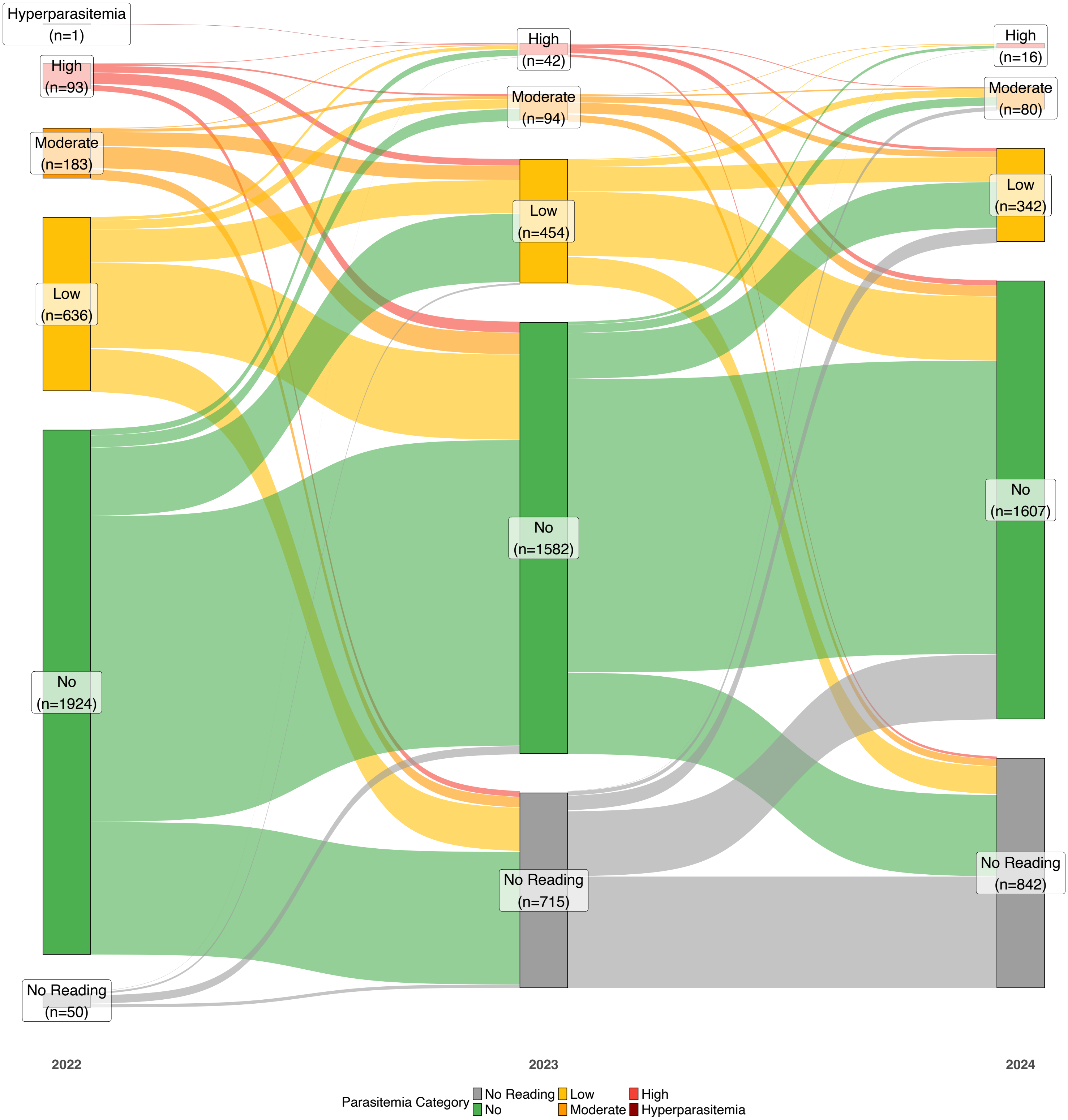


Figure S14: Sankey diagram illustrating the longitudinal dynamics of malaria parasitemia status for participants followed from 2022 to 2024.

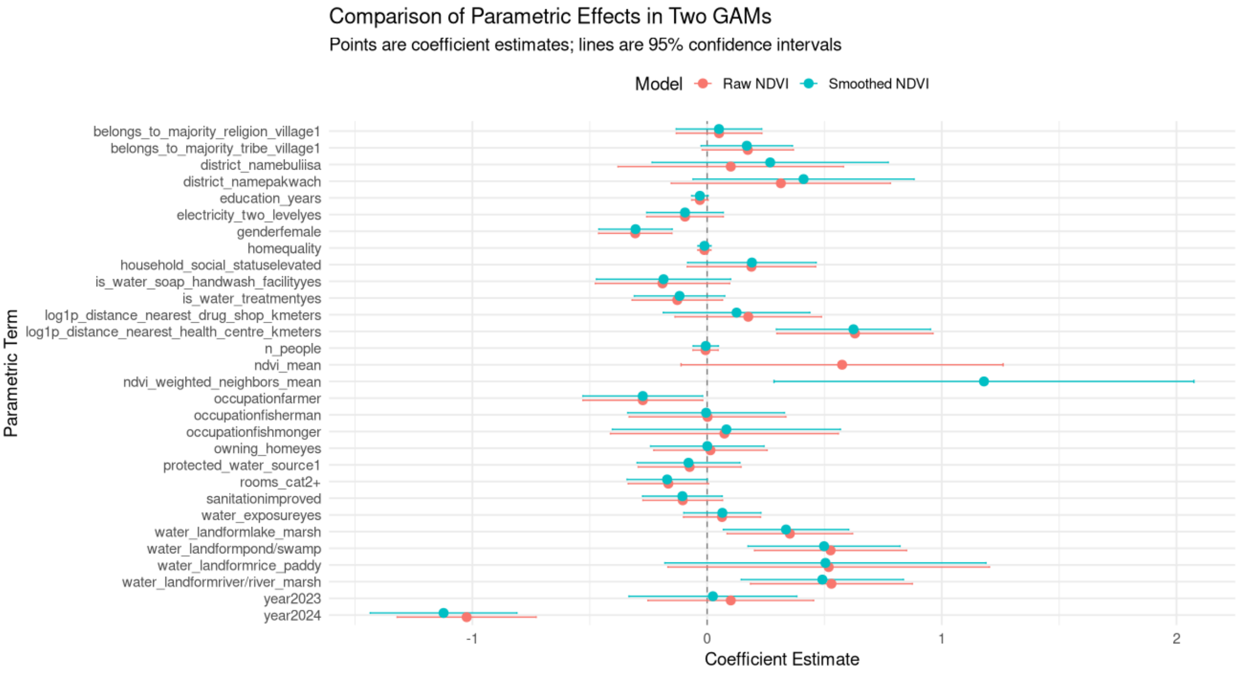


Figure S15: Comparison of model coefficient estimates for models using raw, unsmoothed NDVI (red) versus spatially smoothed NDVI (teal).

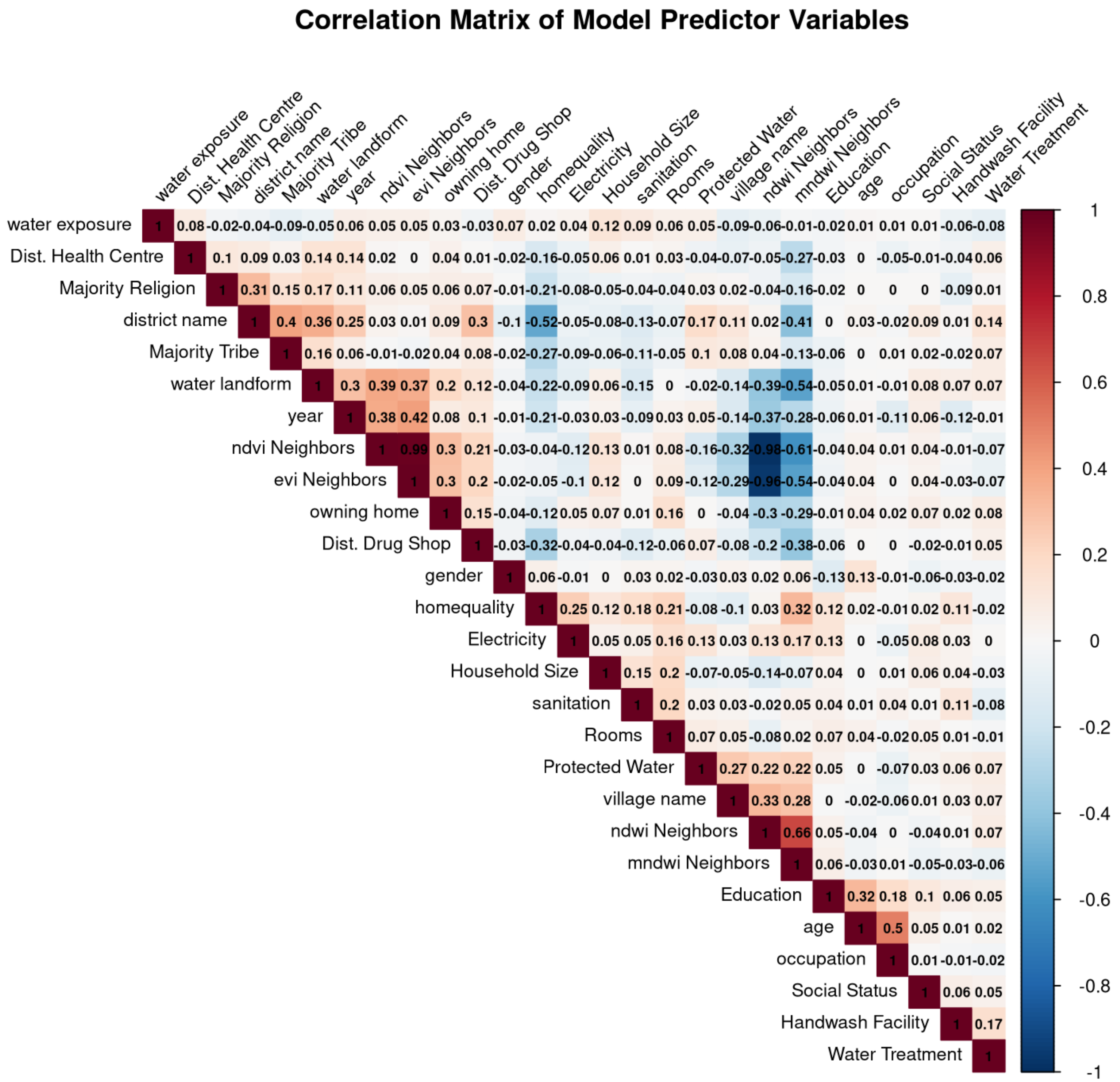


Figure S16: Pairwise Spearman's correlation matrix of all candidate predictor variables.

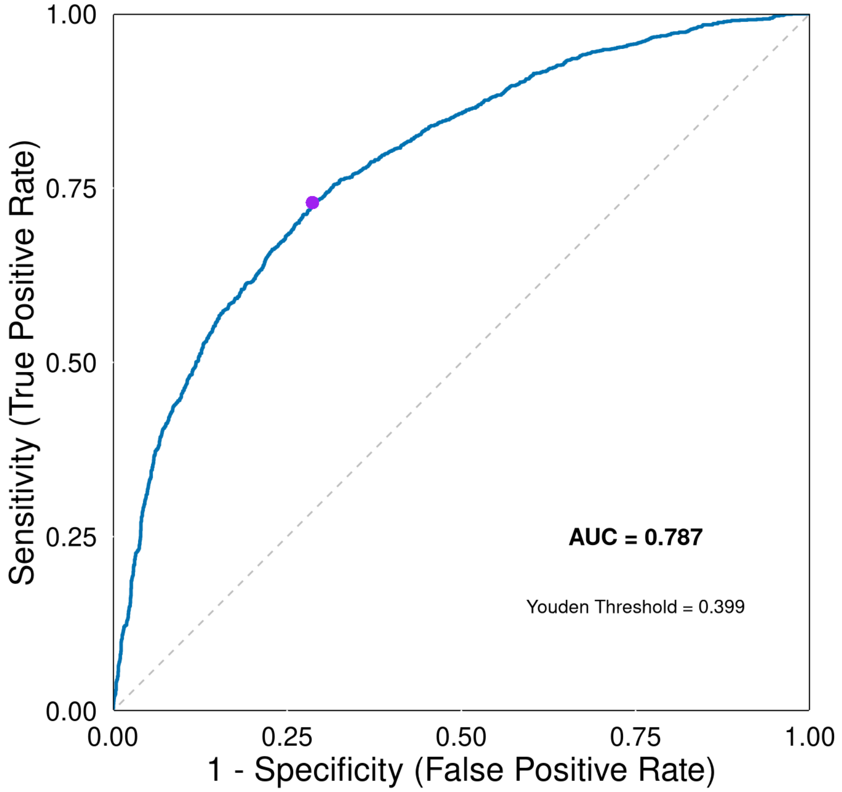


Figure S17: Receiver operating characteristic (ROC) curve for the final malaria prediction model. Area under the curve (AUC) = 0.787.

#### Supplementary Tables

Table S1: Comprehensive summary of Sentinel-2 data availability and quality. Each analysis period spans from December 1 to January 31. Despite high average cloud cover in the source imagery, the median composite method produced a cloud-free mosaic with no excluded pixels.

| District | Analysis Period | Available Scenes | Cloud Cover (%) | Total Pixels | Pixels Used | Excl. Pixels |
|---|---|---|---|---|---|---|
| Pakwach | 2021–2022 | 23 | 43.10 | 2915941 | 2915941 | 0 |
| Buliisa | 2021–2022 | 48 | 43.45 | 3099069 | 3099069 | 0 |
| Mayuge | 2021–2022 | 9 | 58.19 | 5151172 | 5151172 | 0 |
| Pakwach | 2022–2023 | 22 | 30.88 | 2915941 | 2915941 | 0 |
| Buliisa | 2022–2023 | 44 | 36.26 | 3099069 | 3099069 | 0 |
| Mayuge | 2022–2023 | 12 | 55.74 | 5151172 | 5151172 | 0 |
| Pakwach | 2023–2024 | 24 | 43.76 | 2915941 | 2915941 | 0 |
| Buliisa | 2023–2024 | 48 | 48.62 | 3099069 | 3099069 | 0 |
| Mayuge | 2023–2024 | 11 | 58.77 | 5151172 | 5151172 | 0 |

Table S2: Environmental indices by water body type at malacological survey sites.

| Index | | Overall ($n = 437$) | Lake marsh ($n = 162$) | Lake beach ($n = 151$) | Pond/swamp ($n = 60$) | Rice paddy ($n = 9$) | River/marsh ($n = 55$) |
|---|---|---|---|---|---|---|---|
| **NDVI** | | | | | | | |
| | Mean | 0.41 | 0.48 | 0.20 | 0.57 | 0.63 | 0.57 |
| | SD | 0.22 | 0.15 | 0.14 | 0.18 | 0.05 | 0.14 |
| | Median | 0.44 | 0.51 | 0.17 | 0.64 | 0.63 | 0.61 |
| | IQR | [0.22, 0.61] | [0.37, 0.61] | [0.09, 0.28] | [0.52, 0.69] | [0.60, 0.66] | [0.51, 0.68] |
| **EVI** | | | | | | | |
| | Mean | 0.26 | 0.30 | 0.13 | 0.37 | 0.39 | 0.36 |
| | SD | 0.14 | 0.10 | 0.08 | 0.12 | 0.04 | 0.10 |
| | Median | 0.27 | 0.32 | 0.12 | 0.41 | 0.39 | 0.39 |
| | IQR | [0.14, 0.38] | [0.23, 0.38] | [0.07, 0.19] | [0.33, 0.45] | [0.36, 0.41] | [0.28, 0.44] |
| **NDWI** | | | | | | | |
| | Mean | −0.36 | −0.42 | −0.17 | −0.52 | −0.57 | −0.49 |
| | SD | 0.20 | 0.14 | 0.13 | 0.15 | 0.04 | 0.13 |
| | Median | −0.39 | −0.46 | −0.16 | −0.58 | −0.58 | −0.54 |
| | IQR | [−0.55, −0.19] | [−0.54, −0.32] | [−0.24, −0.08] | [−0.60, −0.49] | [−0.61, −0.57] | [−0.58, −0.44] |
| **MNDWI** | | | | | | | |
| | Mean | −0.25 | −0.30 | −0.12 | −0.38 | −0.42 | −0.33 |
| | SD | 0.16 | 0.13 | 0.13 | 0.11 | 0.06 | 0.10 |
| | Median | −0.28 | −0.32 | −0.13 | −0.41 | −0.44 | −0.36 |
| | IQR | [−0.39, −0.14] | [−0.40, −0.23] | [−0.20, −0.06] | [−0.46, −0.37] | [−0.47, −0.38] | [−0.40, −0.29] |

Table S3: Household occupancy by number of rooms.

| Number of Rooms | Number of People Mean (SD) | Median | IQR | Sample Size (n) |
|---|---|---|---|---|
| 1 | 3.2 (1.1) | 3.0 | [3, 4] | 1,286 |
| 2 | 3.7 (1.4) | 3.0 | [3, 5] | 1,878 |
| 3 | 4.0 (1.7) | 4.0 | [3, 5] | 703 |
| 4 | 4.4 (1.7) | 4.0 | [3, 5] | 269 |
| 5 | 4.1 (1.6) | 4.0 | [3, 5] | 82 |
| 6+ | 4.1 (1.7) | 4.0 | [3, 5] | 90 |
| Total | | | | 4,308 |

Table S4: Proportion of RDT-positive participants reporting fever within the past month by antimalarial treatment status among 1431 individuals from 2023-2024, stratified by age group.

| Age Group | | Antimalarial Status Treated | Not Treated | *p*-value |
|---|---|---|---|---|
| 5 to <11 years | Fever | 57 (34.1%) | 24 (25.0%) | 0.160 |
| | No Fever | 110 (65.9%) | 72 (75.0%) | |
| 11 to <20 years | Fever | 25 (25.5%) | 17 (18.1%) | 0.285 |
| | No Fever | 73 (74.5%) | 77 (81.9%) | |
| ≥20 years | Fever | 4 (5.6%) | 16 (14.8%) | 0.096 |
| | No Fever | 67 (94.4%) | 92 (85.2%) | |

Table S5: Effect of occupation on malaria infection with and without adjustment for age and gender.

| Model | Variable | Estimate | SE | z value | p-value |
|---|---|---|---|---|---|
| *Model 1: Occupation only (Deviance explained = 11.3%)* | | | | | |
| | Intercept | −0.062 | 0.100 | −0.619 | 0.536 |
| | Farmer | −1.458 | 0.106 | −13.704 | $< 0.001$ |
| | Fisherman | −1.032 | 0.141 | −7.315 | $< 0.001$ |
| | Fishmonger | −1.270 | 0.231 | −5.495 | $< 0.001$ |
| *Model 2: Occupation + Age (Deviance explained = 19.8%)* | | | | | |
| | Intercept | −0.435 | 0.115 | −3.785 | $< 0.001$ |
| | Farmer | −0.205 | 0.128 | −1.609 | 0.108 |
| | Fisherman | 0.248 | 0.160 | 1.550 | 0.121 |
| | Fishmonger | 0.038 | 0.243 | 0.155 | 0.877 |
| | Age (edf = 6.3) | – | – | – | $< 0.001$ |
| *Model 3: Occupation + Gender (Deviance explained = 12.3%)* | | | | | |
| | Intercept | 0.230 | 0.106 | 2.164 | 0.031 |
| | Female | −0.547 | 0.071 | −7.704 | $< 0.001$ |
| | Farmer | −1.360 | 0.108 | −12.635 | $< 0.001$ |
| | Fisherman | −1.316 | 0.146 | −9.014 | $< 0.001$ |
| | Fishmonger | −1.091 | 0.232 | −4.697 | $< 0.001$ |
| *Model 4: Occupation + Age + Gender (Deviance explained = 20.0%)* | | | | | |
| | Intercept | −0.276 | 0.122 | −2.262 | 0.024 |
| | Female | −0.279 | 0.077 | −3.652 | $< 0.001$ |
| | Farmer | −0.189 | 0.128 | −1.477 | 0.140 |
| | Fisherman | 0.064 | 0.167 | 0.386 | 0.700 |
| | Fishmonger | 0.092 | 0.244 | 0.379 | 0.705 |
| | Age (edf = 6.1) | – | – | – | $< 0.001$ |

Note: All models include village random effects. Reference category for occupation is none/other.

Table S6: Cross-validated confusion matrix and performance metrics for the final malaria prediction model.

| | Reference | | |
|---|---|---|---|
| Predicted | Negative (0) | Positive (1) | Total |
| Negative (0) | 1807 | 481 | 2288 |
| Positive (1) | 725 | 1295 | 2020 |
| Total | 2,532 | 1,776 | 4,308 |

| Performance Metric | Value (95% CI) |
|---|---|
| Accuracy | 0.7201 |
| Sensitivity | 0.7292 |
| Specificity | 0.7137 |
| Positive Predictive Value | 0.6411 |
| Negative Predictive Value | 0.7898 |
| Balanced Accuracy | 0.7214 |
| Cohen's Kappa | 0.4339 |

Table S7: Comparison of Intra-class Correlation Coefficient (ICC) Across Nested Models.

| **Model** | $N_{Obs}$ | $N_{Villages}$ | **Village Var.** ($\sigma^2$) | **ICC** |
|---|---|---|---|---|
| Intercept + Village Random Effect | 4,308 | 52 | 0.3087 | 0.0858 |
| Base Model + Village Random Effect | 4,308 | 52 | 0.2543 | 0.0717 |
| NDVI Only + Village Random Effect | 4,308 | 52 | 0.2420 | 0.0685 |
| MNDWI Only + Village Random Effect | 4,308 | 52 | 0.1008 | 0.0297 |
| Base + NDVI + Village Random Effect | 4,308 | 52 | 0.2081 | 0.0595 |
| Base + MNDWI + Village Random Effect | 4,308 | 52 | 0.1664 | 0.0482 |
| Base + NDVI + MNDWI + Village Random Effect | 4,308 | 52 | 0.1556 | 0.0452 |
| **Main Model (All Predictors)** | **4,308** | **52** | **0.1371** | **0.0400** |

[14] Grace Wahba. *Spline models for observational data.* SIAM, Philadelphia, 1990.
